# Cortical and subcortical connections change after repetitive transcranial magnetic stimulation therapy in cocaine use disorder and predict clinical outcome

**DOI:** 10.1101/2022.09.29.22280253

**Authors:** Jalil Rasgado-Toledo, Victor Issa-Garcia, Ruth Alcalá-Lozano, Eduardo A. Garza-Villarreal, Gabriel González-Escamilla

**Author notes:** **Corresponding:** Eduardo A. Garza-Villarreal, MD, PhD, Assistant Professor, Instituto de Neurobiología, Laboratorio B-03, Universidad Nacional Autonoma de Mexico (UNAM) campus Juriquilla, Boulevard Juriquilla 3001, Santiago de Querétaro, Querétaro, México, C.P. 76230, Gabriel González-Escamilla, PhD, Department of Neurology, Focus Program Translational Neuroscience (FTN), Rhine-Main Neuroscience, Network (rmn²), University Medical Center of the Johannes Gutenberg University Mainz, Langenbeckstrasse 1, 55131 Mainz, Germany, Ruth Alcalá-Lozano, MD, MSc, Psychiatrist and Clinical Research., Instituto Nacional de Psiquiatría Ramon de la Fuente Muñiz. Laboratorio de Neuromodulación., Subdireccion de Investigaciones Clínicas., Calzada México Xochimilco 101. Colonia San Lorenzo Huipulco, C.P. 14370. These authors contributed equally (Jalil Rasgado-Toledo, Victor Issa-Garcia). Shared senior authorship (Eduardo A. Garza-Villarreal, Gabriel González-Escamilla).

## Abstract

**Background:** Cocaine use disorder (CUD) is a worldwide public health condition which is suggested to induce pathological changes in macro- and microstructure. Repetitive transcranial magnetic stimulation (rTMS) has gained attention to induce a reduction in CUD symptoms. Here, we sought to elucidate whether rTMS induces changes on white-matter (WM) microstructure in frontostriatal circuits after two weeks of therapy in patients with CUD, and to test whether baseline WM microstructure of the same circuits has an effect on clinical improvement. This study consisted of a 2-week, parallel group, double-blind, randomized controlled clinical trial (acute phase) (sham [n=23] and active [n=27]), in which patients received two daily sessions of rTMS on the left dorsolateral prefrontal cortex (lDLPFC) as an add-on treatment. T1-weighted and HARDI-DWI at baseline and two weeks after served to evaluate WM microstructure. After active rTMS, results showed a significant increase in neurite density compared to sham rTMS in WM-tracts connecting left DLPFC with left and right vmPFC. Similarly, rTMS showed reduction in orientation dispersion in WM tracts connecting left DLPFC with left caudate nucleus, left thalamus and left vmPFC. Results also showed a greater reduction in craving VAS after rTMS when baseline ICVF was low in WM tracts connecting left caudate nucleus with substantia nigra, left pallidum, and left thalamus with substantia nigra and left pallidum. Our results evidence rTMS-induced WM microstructural changes in fronto-striato-thalamic circuits and support its efficacy as a therapeutic tool in the treatment of CUD. Further, individual clinical improvement may rely on the patient’s individual structural connectivity integrity.

**Highlights:**

- White matter microstructural changes between fronto-striato-thalamic regions after 2 weeks of rTMS.
- Whether rTMS would induce microstructural changes may depend on the baseline integrity of the connections between the striatum, thalamus, and the substantia nigra.
- Our results highlight rTMS as a potential therapeutic tool in the treatment of CUD, due to its ability to modulate altered brain microstructure.

## Introduction

Cocaine use disorder (CUD) is estimated to affect about 20 million people worldwide (global population age ≥15) [1] and to be one of the leading causes of death in young people [2]. CUD induces pathological changes in macrostructure and microstructure of the meso-cortico-limbic system in humans and animal models, possibly due to the changes in synapses and cells themselves [3], which some studies suggest may be reversible [4,5]. Despite the improvement in the treatment of CUD patients in recent years, patients with psychosocial and pharmacological approaches achieve only low to moderate effects on clinical symptomatology such as craving, which may lead to a relapse in substance use [6]. Novel non-invasive interventions such as repetitive transcranial magnetic stimulation (rTMS) have gained attention by attaining positive outcomes in substance use disorders (SUDs) [7]. Although the main action mechanism of rTMS remains to be fully elucidated, the stimulation seems to induce acute modulation of the dysregulated frontostriatal circuits, and it is hypothesized that long treatments may produce neuroplastic changes on these circuits [7,8]. In this sense, rTMS may help patients in adequately overcoming symptoms such as craving and impulsivity by producing changes in micro and macroscopic brain circuits [8–10]. We recently showed that rTMS at 5-Hz over the left dorsolateral prefrontal cortex (lDLPFC) reduced craving and impulsivity in CUD patients after 2 weeks, and increased functional connectivity between lDLPFC and ventromedial PFC (vmPFC), as well as vmPFC and right Angular gyrus. However, to date there is limited evidence on the extent to which rTMS modulates the microstructure of brain circuits in CUD.

Magnetic resonance imaging (MRI) has open new opportunities for the study of microstructural abnormalities, these include the neurite orientation dispersion and density imaging model (NODDI), which allows to evaluate subtle tissue characteristics by modeling water diffusion within neurites and the free-water around them [11]. Microstructural characterization with NODDI is possible through the use of different metrics: 1) intra-cellular volume fraction or neurite density index (ICVF), which assesses the density of axons and dendrites, and has been proposed as a sensitive marker for myeloarchitecture and axonal degeneration, 2) isotropic volume fraction (ISOVF) which measures the free water of extracellular compartment as an indirect metric for neuroinflammation, and 3) Orientation dispersion index (OD) which quantifies bending, fanning and the orientational configuration of axons that helps to measure axonal organization [12,13]. Using NODDI offers a unique opportunity to understand the microstructural plasticity of the human brain as a result of rTMS treatment for CUD. This may be particularly interesting as NODDI can be developed into an indicator of a state-of-disorder and treatment-outcome biomarker, as has been proposed for other diseases [14–18], and described in context of the state-of-disorder for alcohol and cocaine use disorders, however not for treatment-outcome [19,20].

In this study we wished to evaluate whether rTMS induced changes on white-matter (WM) microstructure of brain circuits related to SUDs after two weeks of rTMS treatment in CUD patients, and to test whether baseline WM microstructure of those circuits could predict clinical improvement. To achieve this, we used longitudinal multi-shell diffusion weighted imaging and NODDI analysis.

## Methods

### Participants

Data was obtained from a clinical trial conducted at the Clinical Research Division of the National Institute of Psychiatry in Mexico City, Mexico that was approved by the Institutional Ethics Research Committee (CEI/C/070/2016) and registered at ClinicalTrials.gov (NCT02986438). During the clinical trial, patients with cocaine use disorder (CUD) who met the inclusion and did not meet exclusion criteria provided written informed consent to participate, per the Declaration Helsinki. Patients were recruited via flyers on substance use disorder (SUD) clinics and medical institutes in the metropolitan area of Mexico City and through advertisements in social media. The clinical trial consisted of a 2-week, parallel group, double-blind, randomized controlled trial (acute phase) followed by a 6-month open-label trial

(maintenance phase). Repetitive transcranial magnetic stimulation (rTMS) was delivered over the left dorsolateral prefrontal cortex (DLPFC) as an add-on to standard treatment (full information can be found in supplementary material). Clinical, behavioral and MRI data was acquired at baseline (T0), 2 weeks (T1), 3 months (T2) and 6 months (T3). The main results of the clinical trial and functional analysis have been published [21]. For the purpose of this new study, we focused on the double-blind 2-week acute phase (T0-T1) and the relationship between clinical improvement and WM structure measured using a novel HARDI-DWI sequence and NODDI analysis. We analyzed 50 participants (sham: 23, active: 27) and the overview of demographic data is shown in **Table 1**. Detailed information about recruitment, CONSORT flowchart, and a complete demographics table, as well as the main clinical and fMRI study results can be found in our previous publication by Garza-Villarreal et al. [21] and the Supplementary Material.

**Table 1.** Descriptive statistics of demographics, behavioral instruments and NODDI metrics by time point and sham/active groups.

|  | Sham |  | Active |  |
| --- | --- | --- | --- | --- |
|  | T0 (n = 23) | T1 (n = 20) | T0 (n = 30) | T1 (n = 25) |
| **Demographic** |  |  |  |  |
| Sex, n (%) |  |  |  |  |
| Male | 20 (87%) | - | 25 (83.3%) | - |
| Female | 3 (13%) | - | 5 (16.7%) | - |
| Age in years, mean ± SD | 33.3 ± 8.1 | - | 36.1 ± 6.8 | - |

|  |  |  |  |  |
| --- | --- | --- | --- | --- |
| Years of education, mean ± SD | 13.4 ± 2.8 | - | 12.9 ± 3.1 | - |
| **Behavioral instrument, mean ± SD** |  |  |  |  |
| Craving: VAS | 2.9 ± 2.9 | 2.3 ± 2.5 | 3.7 ± 3.5 | 1.5 ± 2.4 |
| Craving: CCQ-now | 146 ± 47.8 | 129.2 ± 46.2 | 148.7 ± 51.5 | 114.8 ± 44.7 |
| Impulsivity: BIS11 | 62.6 ± 16.9 | 59.8 ± 21.4 | 65 ± 16.6 | 53.4 ± 17.1 |
| **NODDI metrics, mean ± SD** | T0 (n = 23) | T1 (n = 20) | (n = 27) | (n = 23) |
| ICVF: Caud2Medulla | 0.812 ± 0.056 | 0.814 ± 0.063 | 0.813 ± 0.053 | 0.825 ± 0.066 |
| ICVF: Caud2Palli | 0.818 ± 0.061 | 0.815 ± 0.066 | 0.818 ± 0.056 | 0.823 ± 0.065 |
| ICVF: DLPFC2Caud | 0.631 ± 0.076 | 0.624 ± 0.073 | 0.627 ± 0.051 | 0.639 ± 0.063 |
| ICVF: DLPFC2vmPFC | 0.515 ± 0.129 | 0.477 ± 0.1 | 0.451 ± 0.116 | 0.484 ± 0.126 |
| ICVF: DLPFC2Thal | 0.676 ± 0.068 | 0.673 ± 0.071 | 0.676 ± 0.038 | 0.686 ± 0.056 |
| ICVF: rAngG2rDLPFC | 0.663 ± 0.066 | 0.668 ± 0.073 | 0.674 ± 0.036 | 0.674 ± 0.045 |
| ICVF: Thal2Medulla | 0.757 ± 0.08 | 0.761 ± 0.081 | 0.759 ± 0.06 | 0.773 ± 0.072 |
| ICVF: Thal2Palli | 0.771 ± 0.067 | 0.775 ± 0.07 | 0.776 ± 0.058 | 0.784 ± 0.066 |
| ICVF: vmPFC2DLPFC | 0.476 ± 0.126 | 0.441 ± 0.1 | 0.422 ± 0.135 | 0.461 ± 0.142 |
| ISOVF: Caud2Medulla | 0.298 ± 0.068 | 0.296 ± 0.077 | 0.336 ± 0.072 | 0.333 ± 0.085 |
| ISOVF: Caud2Palli | 0.212 ± 0.064 | 0.214 ± 0.07 | 0.247 ± 0.067 | 0.249 ± 0.076 |
| ISOVF: DLPFC2Caud | 0.178 ± 0.057 | 0.175 ± 0.059 | 0.192 ± 0.042 | 0.192 ± 0.04 |
| ISOVF: DLPFC2vmPFC | 0.288 ± 0.078 | 0.287 ± 0.086 | 0.281 ± 0.084 | 0.283 ± 0.086 |
| ISOVF: DLPFC2Thal | 0.219 ± 0.055 | 0.217 ± 0.061 | 0.242 ± 0.046 | 0.242 ± 0.05 |
| ISOVF: rAngG2rDLPFC | 0.143 ± 0.038 | 0.149 ± 0.043 | 0.164 ± 0.032 | 0.163 ± 0.035 |
| ISOVF: Thal2Medulla | 0.393 ± 0.09 | 0.388 ± 0.101 | 0.434 ± 0.071 | 0.431 ± 0.081 |
| ISOVF: Thal2Palli | 0.313 ± 0.079 | 0.313 ± 0.086 | 0.348 ± 0.075 | 0.349 ± 0.082 |
| ISOVF: vmPFC2DLPFC | 0.144 ± 0.047 | 0.139 ± 0.049 | 0.138 ± 0.048 | 0.147 ± 0.049 |
| OD: Caud2Medulla | 0.378 ± 0.041 | 0.373 ± 0.047 | 0.398 ± 0.044 | 0.393 ± 0.052 |
| OD: Caud2Palli | 0.355 ± 0.028 | 0.351 ± 0.028 | 0.362 ± 0.033 | 0.362 ± 0.037 |
| OD: DLPFC2Caud | 0.423 ± 0.041 | 0.411 ± 0.031 | 0.41 ± 0.037 | 0.416 ± 0.035 |
| OD: DLPFC2vmPFC | 0.399 ± 0.073 | 0.391 ± 0.083 | 0.365 ± 0.093 | 0.381 ± 0.094 |
| OD: DLPFC2Thal | 0.421 ± 0.037 | 0.411 ± 0.03 | 0.418 ± 0.03 | 0.42 ± 0.027 |
| OD: rAngG2rDLPFC | 0.415 ± 0.024 | 0.405 ± 0.019 | 0.413 ± 0.027 | 0.411 ± 0.024 |
| OD: Thal2Medulla | 0.429 ± 0.064 | 0.423 ± 0.072 | 0.455 ± 0.049 | 0.446 ± 0.053 |
| OD: Thal2Palli | 0.413 ± 0.04 | 0.407 ± 0.044 | 0.426 ± 0.034 | 0.423 ± 0.037 |

|  |  |  |  |  |
| --- | --- | --- | --- | --- |
| OD: vmPFC2DLPFC | 0.347 ± 0.093 | 0.327 ± 0.089 | 0.294 ± 0.095 | 0.329 ± 0.096 |
| Mean (standard deviation) for demographics and NODDI metrics for each tract: n = sample size, T0 = baseline, T1 = after 2 week; ICVF = intra-cellular volume fraction, ISOVF = isotropic volume fraction, OD = Orientation dispersion: WM-ROI tracts. |  |  |  |  |

### Clinical instruments

**Table 1** shows descriptive statistics on the scores of the clinical instruments. A full list with their descriptions, and mean scores can be found in Garza-Villarreal et al. [21]. Here, we limit to describe the following which were employed in the analyses.

#### Cocaine Craving Visual Analog Scale (VAS)

We designed a VAS to evaluate the participants’ craving at the moment of the assessment. The VAS consisted of a continuous 10 cm line (including 2 decimals) in which the left point at 0 cm meant “no craving” and the right point at 10 cm meant “the most intense craving” [22]. The participants were asked to mark with a cross the intensity of their craving at the moment of assessment.

#### Cocaine Craving Questionnaire Now (CCQ-now)

The CCQ-Now is a questionnaire containing 45 items that explore cocaine craving at the moment of assessment [23]. The items are related to: desire to use cocaine, intention and planning to use cocaine, anticipation of positive outcome, anticipation of relief from withdrawal or dysphoria and lack of control overuse. The higher the score is, the higher current cocaine craving. We employed the CCQ-now Spanish translation which was validated in the Mexican population [24].

#### Barratt Impulsivity Scale Version 11 (BIS-11)

The BIS is more than 50 years old [25] and has been extensively revised into the BIS-11 [26]. This version of the scale consists of 30 items that describe impulsive and non-impulsive behaviors related to 3 main categories: attentional, motor, and non-planning impulsiveness. The higher the total score is, the higher the impulsivity of the participant. We used the Spanish translation of the BIS-11 [27].

### Transcranial magnetic stimulation

During the acute phase, a MagPro R30+Option magnetic stimulator and an eight-shaped B65-A/P coil (MagVenture, Alpharetta, GA) was used to deliver active or sham rTMS over the left DLPFC. The orientation of the coil was anterior-medial and the position was determined using either the 5.5 cm anatomic criterion or the Beam F3 method. Stimulation protocol consisted of 10 weekdays of 2 sessions with 50 trains of 50 pulses at 5 Hz at 100% motor threshold. Each train had a 10-second inter-train interval and each session a 15-minute inter-session interval. A total of 5,000 pulses were delivered per day. For all details on the rTMS protocol please refer to the Supplementary Materials.

### Simulation of TMS electrical field

We used SimNIBS [28] version 3.2.3 to perform simulations of the TMS electrical field generated on the participants cortex. We reconstructed head meshes from T1w images using *headreco [29]*. During the study, Brain Navigator was not available, hence, a vitamin E capsule fiducial was used to identify the stimulation target. These scalp coordinates were projected into the cortex and converted to MNI coordinates reported in and with a procedure explained in detail in Garza-Villarreal et al. [21]. We converted these MNI coordinates into the native space by using SimNIBS’s *mni2subjects_coords* function which were projected back into the scalp before running the subjects’ simulations. Simulations were run with the corresponding coordinates, coil orientation (anterior-medial) and 100% motor threshold. For the simulation, we used the MagVenture_MC_B70 (MCF-B70) coil, since there are no differences in the induced field between the B65-A/P and the MCF-B70 coil. Electric field norms from simulations were mapped into *fsaverage* and then averaged across participants to obtain **Figure 1**.

**Figure 1.**
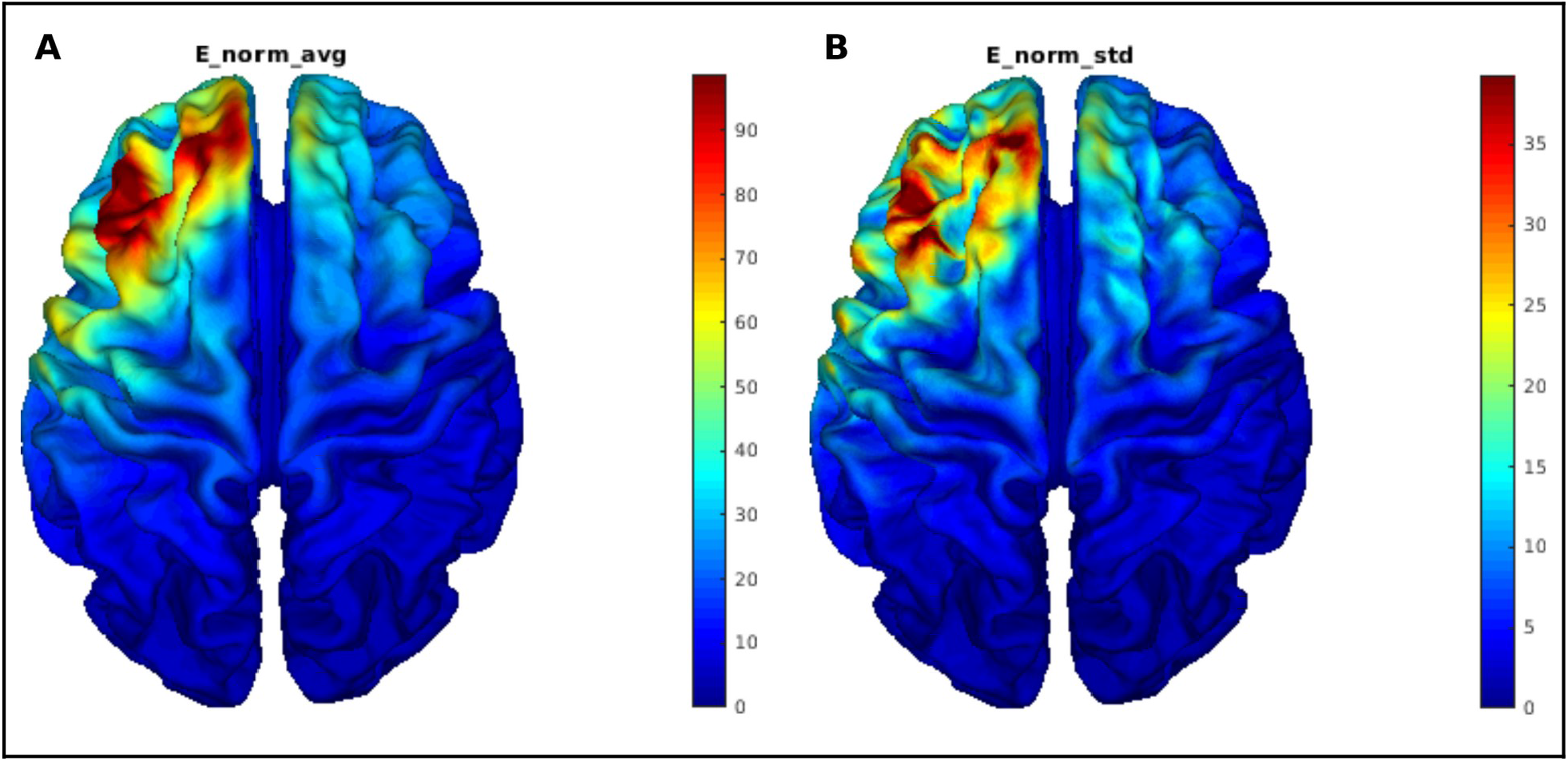
Visualization of A) mean (V/m) and B) standard deviation (V/m) of electric field norms from simulations across subjects.

### Magnetic resonance imaging acquisition

T1-weighted (T1w) and High angular resolution diffusion-weighted imaging (DWI-HARDI) sequences were acquired using a Philips Ingenia 3T scanner (Philips, USA) with a 32-channel Philips head coil. Parameters of T1w 3D FFE sagittal images were: TR/TE = 7/3.5ms, FA = 8 degrees, FOV = 240mm2, matrix = 240×240, voxel = 1×1×1mm, gap = 0. DWI-HARDI used a spin echo (SE) sequence, TR/TE =9000/127ms, FOV=230mm2 (for 4 participants=224mm2), matrix=96×96 (for 4 participants=112×112), number of slices=57 (for 4 participants=58), gap=0, plane=axial, voxel=2.4×2.4×2.5 mm (for 4 participants=2×2×2.3 mm), directions: 8=b0, 32=b-value 1,000s/mm2 and 96=b-value 2,500s/mm2 (for 4 participants: 96=b-value 3,000s/mm2), total=136 directions. Field maps intended for DWI-HARDI use were acquired using a SE-EPI sequence with the following parameters: TR/TE=9000/127ms, flip angle=90°, matrix=128×128 (for 4 participants=112×112), voxel size=1.8×1.8×2.5mm (for 4 participants=2×2×2.3mm), number of slices=57 (for 4 participants=58), phase encoding direction=PA. A total of 7 b0 volumes were acquired.

### Preprocessing

T1w preprocessing was performed using FMRIPREP v1.5.5 [30][RRID:SCR_016216], a Nipype [31] [RRID:SCR_002502] based tool. Each T1w volume was corrected for INU (intensity non-uniformity) using N4BiasFieldCorrection v2.1.0 [32] and skull-stripped using *antsBrainExtraction*.*sh* v2.1.0 (using the OASIS template). Brain surfaces were reconstructed using *recon-all* from FreeSurfer v6.0.1 [33] [RRID:SCR_001847] and the brain mask estimated previously was refined with a custom variation of the method to reconcile ANTs-derived and FreeSurfer-derived segmentations of the cortical gray-matter (GM) of Mindboggle [34][RRID:SCR_002438].

DWI-HARDI preprocessing was performed in MRtrix version 3.0_RC3 [35], using an established pipeline [36] which included noise removal by principal component analysis (PCA) using the Marchenko-Pastur (MP) universal distribution [37], followed by removal of Gibbs ringing artifacts using the method of local subvoxel-shifts [38]. Afterwards, we performed a correction for Eddy currents distortions and motion artifacts via FSL [39]. Data was collected with reversed phase-encoded blips, resulting in pairs of images with distortions going in opposite directions (PA). Mean PA field maps were downsampled using FSL *flirt [40–42]* with an identity matrix and trilinear interpolation to match the voxel size of the mean AP b0 from DWI-HARDI images. From these pairs the susceptibility-induced off-resonance field was estimated using a method similar to that described in Andersson *et al. [43]* as implemented in FSL [44] and the two images were combined into a single corrected one. A brain mask was obtained using *dwi2mask [45]* at this point. Finally, a bias field correction was performed [32,46].

### Processing

All b0 volumes were extracted from preprocessed HARDI images, which were then averaged into a single mean AP b0 volume. Coregistration of this mean volume from each participant to their anatomical T1w image was conducted using Freesurfer’s *bbregister [42]* to obtain a .*dat* registration file and then to apply it to the preprocessed HARDI image using *mri_vol2vol* with trilinear interpolation and to the brain mask using the same function with nearest neighbor interpolation. We used AMICO’s v1.2.10 [47] implementation of NODDI [13] to fit the model and extract its metrics (ICVF, ISOVF, and OD) within the brain mask voxels.

### White matter regions of interest

We chose white matter (WM) regions of interest (ROI) corresponding to frontostriatal circuits based on a reported pathological brain target [21]. These circuits connects the following GM regions: left dorsolateral prefrontal cortex (DLPFC), left caudate nucleus, substantia nigra, left globus pallidus, left thalamus, left and right ventromedial prefrontal cortex (vmPFC), and right angular gyrus (AngG). To select ROI-coordinates, we used the IIT Human Brain Atlas version 5.0 [48,49] to select the following WM ROIs: **1)** left caudate nucleus and axial section through medulla (the closest in atlas to substantia nigra) (*Caud2Medulla*, label 3785), **2)** left caudate nucleus and left pallidum (*Caud2Palli*, label 3739), **3)** left rostral middle frontal cortex (RMFC, the closest in atlas to left DLPFC) and left caudate nucleus (*DLPFC2Caud*, label 2637), **4)** left RMFC and right medial orbitofrontal cortex (MOC, the closest in atlas to vmPFC) (*DLPFC2rvmPFC*, label 2662), **5)** left RMFC and left thalamus (*DLPFC2Thal*, label 2636), **6)** right inferior parietal cortex (IPC, the closest in atlas to right AngG) and right RMFC (*rAngG2rDLPFC*, label 5675), **7)** left thalamus and axial section through medulla (*Thal2Medulla*, label 3685), **8)** left thalamus and left pallidum (*Thal2Palli*, label 3639), **9)** left MOC and left RMFC (*vmPFC2DLPFC*, label 1326).

For each WM-ROI mask, we selected every voxel that was within the top 60 most probable fiber connections passing through that voxel by processing the file *IIT_WM_atlas_256*.*nii*.*gz* with *fslmaths*. For WM-ROI masks 1 and 7 connecting to substantia nigra (axial section through medulla) an inferior manual deletion of all slices (Z ≤ 87) was performed using *fsleyes*, since the WM mask extended inferiorly beyond substantia nigra when overlaying over *IITmean_t1_256*.*nii*.*gz*. A similar process was performed for WM-ROI masks (2 and 8) connecting to globus pallidus to delete all slices (Z ≤ 101), since the WM mask extended inferiorly beyond globus pallidus.

To registering WM-ROI masks into subjects’ native spaces, we first performed a coregistration between Freesurfer’s *brain*.*mgz* and *IITmean_t1_256*.*nii*.*gz* by using *mri_robust_register [50]* to obtain an LTA registration file. The inverse of the LTA file was then used to warp with *mri_vol2vol* the WM-ROI masks into each subject’s native space. These masks in native space were then used to obtain the mean NODDI metrics within each WM-ROI using R version 4.1.0 and *oro*.*nifti* version 0.11.0. **Table 1** shows the descriptive statistics of these NODDI metrics across subjects.

### Statistical analyses

#### 1) Changes in white matter microstructure two weeks after rTMS

Linear mixed-effects models were fitted to elucidate possible effects from active rTMS on frontostriatal WM tracts based on NODDI metrics, while controlling for age and education years. The 27 fitted models were of the following structure:

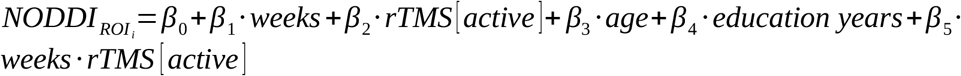

in which *NODDI* _*ROI*_ *i* was the ICVF, ISOVF, or OD of each of the 9 WM-ROI. Supplementaries tables and figures 1-3 depict the estimates of *β*_5_ from these 27 models.

#### 2) Baseline NODDI values as a predictor of clinical response to rTMS after two weeks

To investigate whether baseline NODDI metrics from frontostriatal circuits’ WM tracts could predict clinical response to rTMS, several linear mixed effects models were also fitted. The dependent variable was the clinical instrument score and the independent variables included the NODDI metric of each of the 9 WM-ROI, as shown in the following equation:

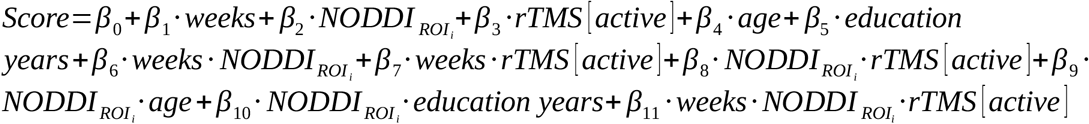

in which *Score* was the score from VAS, CCQ-now, or BIS-11 and *NODDI* _*ROI*_ *i* same as previously defined. Supplementaries tables and figures 4-12 depict the estimates of *β*_11_ from these 81 models (3 NODDI metrics *x* 9 WM-ROI *x* 3 scores from clinical instruments).

We corrected p-values using the false discovery rate or FDR method [51] on each of the 12 hypotheses families, each containing the 9 interactions of interest corresponding to the model of each WM-ROI. All statistical analyses were calculated using R version 4.1.0. Linear mixed models were performed using *lme4, effects, lmerTest* libraries, and *emmeans* for Cohen’s effect size (d) calculated by estimated marginal means (emm).

## Results

### Simulation of TMS electrical field

The simulation of the electric field indicated a maximal average electric field norm (111.46 V/m) in the middle frontal gyrus (**Figure 1**).

### Changes in WM microstructure after 2 weeks rTMS

After 2 weeks, active rTMS showed significant increase in neurite density (ICVF or the density of axons and dendrites) compared to Sham rTMS in WM tracts connecting left DLPFC with left vmPFC (p=0.013, FDR=0.058, d=-0.278; sham-T1: 0.422±0.135, active-T1: 0.461±0.142) and right (p=0.009, FDR=0.058, d=-0.112; sham-T1: 0.451±0.116, active-T1: 0.484±0.126) (**Table 2** and **Figure 2**), though it did not survive multiple comparisons at q=0.05. Similarly, rTMS showed increasing in orientation dispersion (OD or axon organization) in WM tracts connecting left DLPFC with left caudate nucleus (p=0.021, FDR=0.094, d=-0.087; sham-T1: 0.41±0.037, active-T1: 0.416±0.035), left thalamus (p=0.045, FDR=0.135, d=-0.489; sham-T1: 0.418±0.03, active-T1: 0.42±0.027), and left vmPFC (p=0.012, FDR=0.094, d=0.024; sham-T1: 0.294±0.095, active-T1: 0.329±0.096), without surviving multiple comparisons either (**Table 2** and **Figure 2**). Here we show that although all our changes were significant after 2 weeks, none survived our strict multiple comparisons and only the tracts connecting lDLPFC and vmPFC were near the threshold (FDR 5%) with a small effect size increase in density of axons and dendrites after rTMS. The tract connecting lDLPFC and left thalamus showed a medium effect size with an increase in axon organization. We did not find a correlation between changes in WM tracts and craving or impulsivity.

**Table 2.** Coefficients of linear mixed effects models’ interaction (weeks:rTMS[active]) showing the effect of rTMS on NODDI metrics from frontostriatal circuits’ white matter (WM) tracts.

| WM ROI | Estimate | Std. error | df | Effect size | p-value | FDR* |
| --- | --- | --- | --- | --- | --- | --- |
| **Neurite density (ICVF)** |  |  |  |  |  |  |
| DLPFC2rvmpFC | 0.034 | 0.012 | 42.36 | -0.112 | 0.009* | 0.058 |
| vmPFC2DLPFC | 0.038 | 0.015 | 42.31 | -0.278 | 0.013* | 0.058 |
| **Orientation dispersion (OD)** |  |  |  |  |  |  |
| DLPFC2Caud | 0.009 | 0.004 | 40.65 | -0.087 | 0.021* | 0.094 |
| DLPFC2Thal | 0.006 | 0.003 | 40.68 | -0.489 | 0.045* | 0.135 |
| vmPFC2DLPFC | 0.027 | 0.010 | 41.91 | 0.024 | 0.012* | 0.094 |

DLPFC2Caud: WM connecting left rostral middle frontal cortex (RMFC) and left caudate nucleus, DLPFC2rvmpFC: WM connecting left RMFC and right medial orbitofrontal cortex (MOC), DLPFC2Thal: WM connecting left RMFC and left thalamus, vmPFC2DLPFC: WM connecting left MOC and left RMFC. \*False discovery rate (FDR) adjustment was implemented with p-values of their own metric, full values are found in supplementaries tables 1-3.

**Figure 2.**
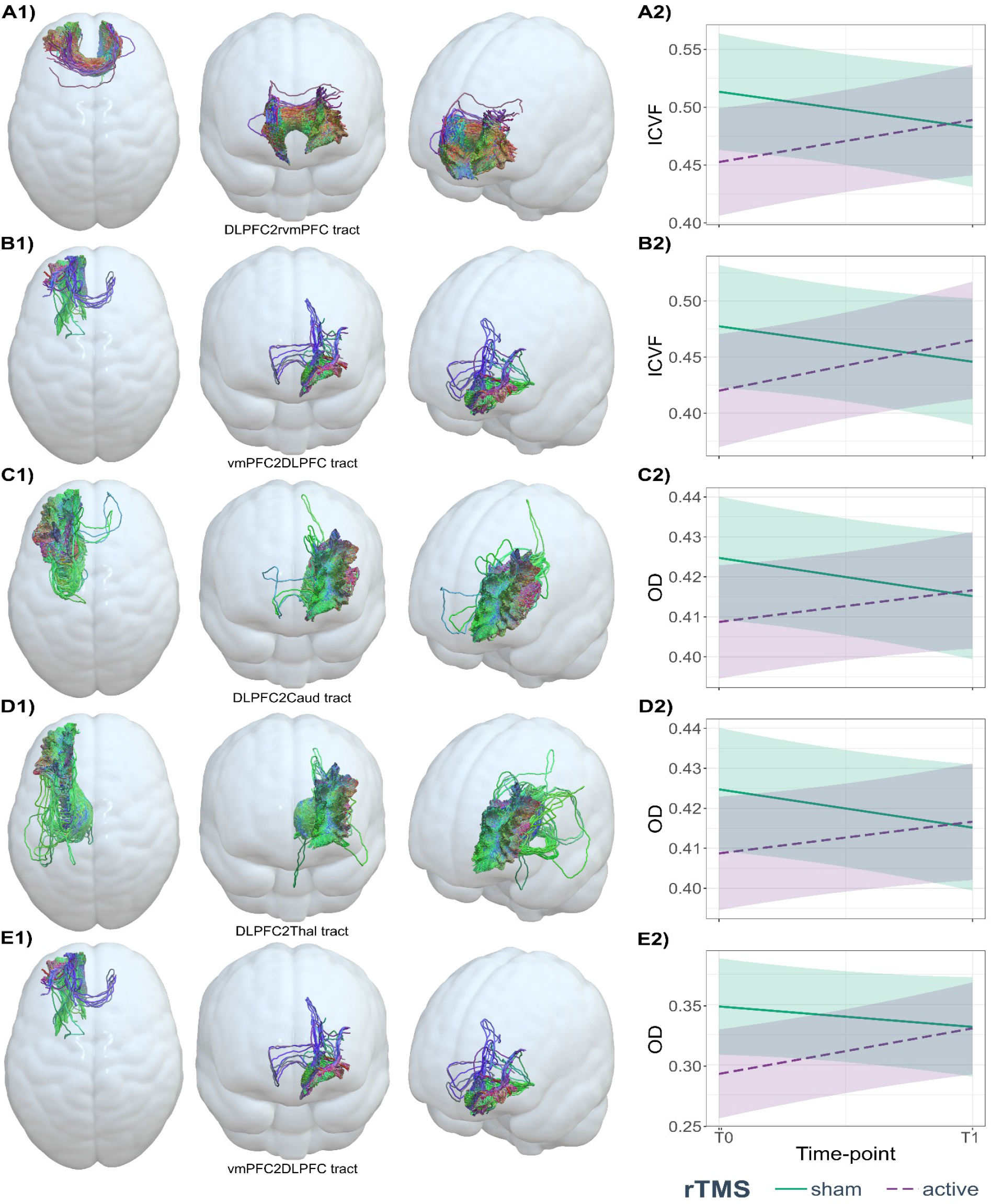
Effect of rTMS on neurite density (ICVF) and orientation dispersion (OD) from frontostriatal circuits’ WM tracts. A2) Mean ICVF values at baseline and after two weeks of the white matter connecting the left rostral middle frontal cortex and right medial orbitofrontal cortex (A1: DLPFC2rvmPFC), B2) Mean ICVF values at baseline and after two weeks of the white matter connecting left medial orbitofrontal cortex and left rostral middle frontal cortex (B1: vmPFC2DLPFC), C2) Mean OD values at baseline and after two weeks of the white matter connecting left rostral middle frontal cortex and left caudate nucleus (C1: DLPFC2Caud), D2) Mean OD values at baseline and after two weeks of the white matter connecting left rostral middle frontal cortex and left thalamus (D1: DLPFC2Thal), E2) Mean OD values at baseline and after two weeks of the white matter connecting left medial orbitofrontal cortex and left rostral middle frontal cortex (E1: vmPFC2DLPFC).

### Baseline NODDI values as a predictor of clinical outcome after rTMS

We found a greater reduction in craving VAS after 2 weeks rTMS when baseline ICVF was low in WM tracts connecting left caudate nucleus with substantia nigra (p=0.007, FDR=0.021, d=0.729) and left pallidum (p=0.004, FDR=0.018, d=0.738), and those connecting left thalamus with substantia nigra (p=0.012, FDR=0.027, d=0.649) and left pallidum (p=0.002, FDR=0.018, d=0.723) (**Table 3** and **Figure 3A**).

**Table 3.** Coefficients of linear mixed effects models’ 3-way interaction (weeks:NODDI:rTMS[active]) predicting decrease in craving VAS/total BIS11 using baseline NODDI metrics from frontostriatal circuits’ WM tracts.

| WM ROI | Estimate | Std. error | df | Effect size | p-value | FDR* |
| --- | --- | --- | --- | --- | --- | --- |
| **ICVF \| Cocaine Craving Visual Analog Scale (VAS)** |  |  |  |  |  |  |
| Caud2Medulla | 22.61 | 7.93 | 40.12 | 0.729 | 0.007* | 0.021* |
| Caud2Palli | 21.13 | 6.95 | 40.17 | 0.738 | 0.004* | 0.018* |
| Thal2Medulla | 16.30 | 6.17 | 40.07 | 0.649 | 0.012* | 0.027* |
| Thal2Palli | 21.92 | 6.61 | 40.13 | 0.723 | 0.002* | 0.018* |
| **ISOVF \| Barratt Impulsivity Scale Version 11 (total BIS-11)** |  |  |  |  |  |  |
| DLPFC2rvmpFC | -49.81 | 23.25 | 39.63 | 1.031 | 0.038* | 0.171 |
| vmPFC2DLPFC | -103.78 | 40.96 | 40.51 | 0.925 | 0.015* | 0.135 |
| **OD \| Barratt Impulsivity Scale Version 11 (total BIS-11)** |  |  |  |  |  |  |
| DLPFC2rvmpFC | -56.76 | 23.43 | 39.80 | 0.827 | 0.020* | 0.180 |

Caud2Medulla: WM connecting left caudate nucleus and axial section through medulla, Caud2Palli: WM connecting left caudate nucleus and left pallidum, DLPFC2rvmpFC: WM connecting left rostral middle frontal cortex (RMFC) and right medial orbitofrontal cortex (MOC), Thal2Medulla: WM connecting left thalamus and axial section through medulla, Thal2Palli: WM connecting left thalamus and left pallidum, vmPFC2DLPFC: WM connecting left MOC and left RMFC. \*False discovery rate (FDR) adjustment was implemented with p-values of their own metric, full values are found in supplementaries tables 4-12.

**Figure 3.**
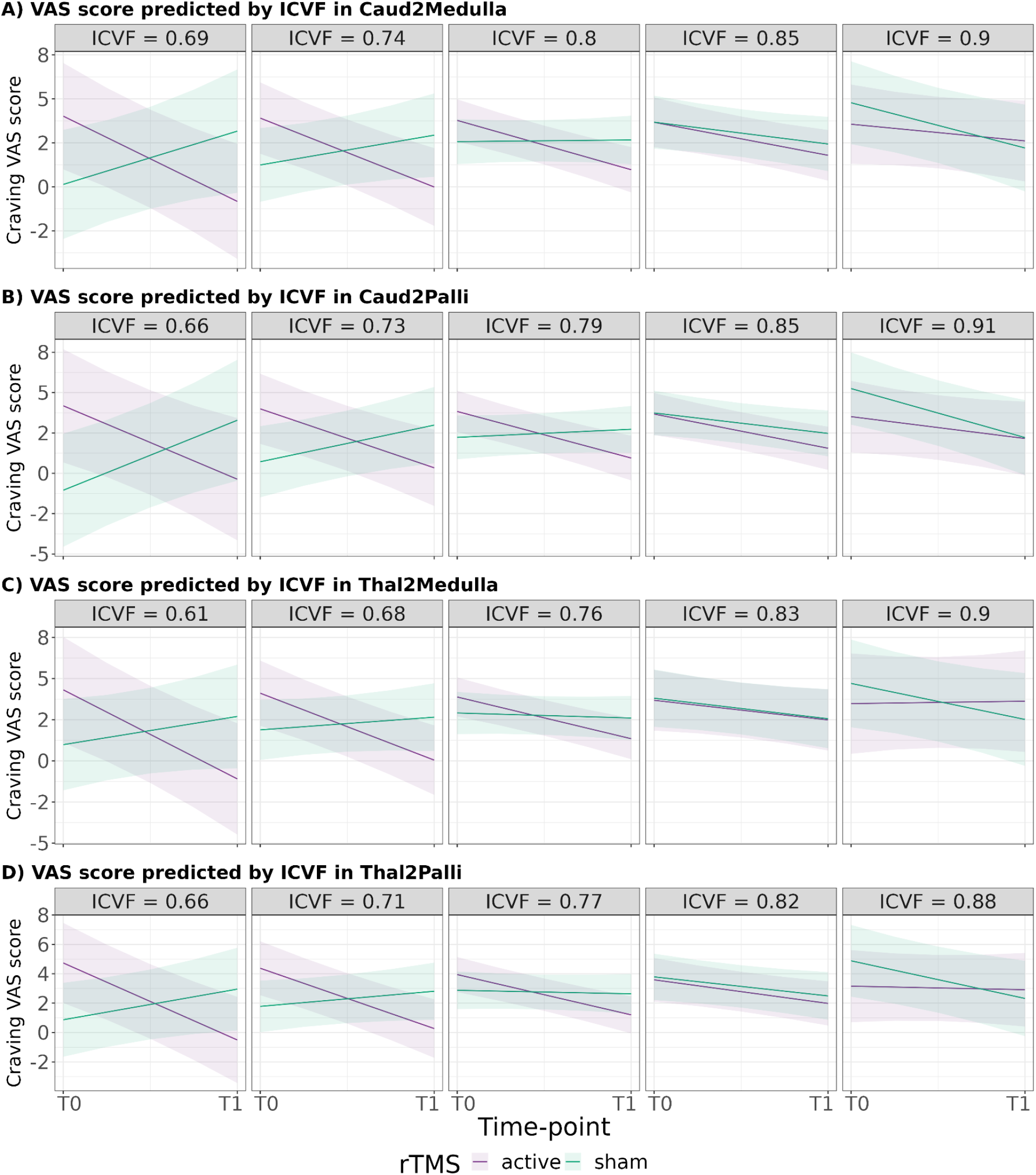
Prediction of craving VAS scores by the effect of rTMS on ICVF from frontostriatal circuits’ WM tracts. A) prediction of craving VAS scores from different ICVF values in WM connecting left caudate nucleus and axial section through medulla (Caud2Medulla), B) prediction of craving VAS scores from different ICVF values in WM connecting left caudate nucleus and left pallidum (Caud2Palli), C) prediction of craving VAS scores from different ICVF values in WM connecting left thalamus and axial section through medulla (Thal2Medulla), D) prediction of craving VAS scores from different ICVF values in WM connecting left thalamus and left pallidum (Thal2Palli). VAS = craving Visual Analog Scale, ICVF = neurite density index, T0 = baseline time-point, T1 = after two weeks of rTMS protocol.

**Figure 4.**
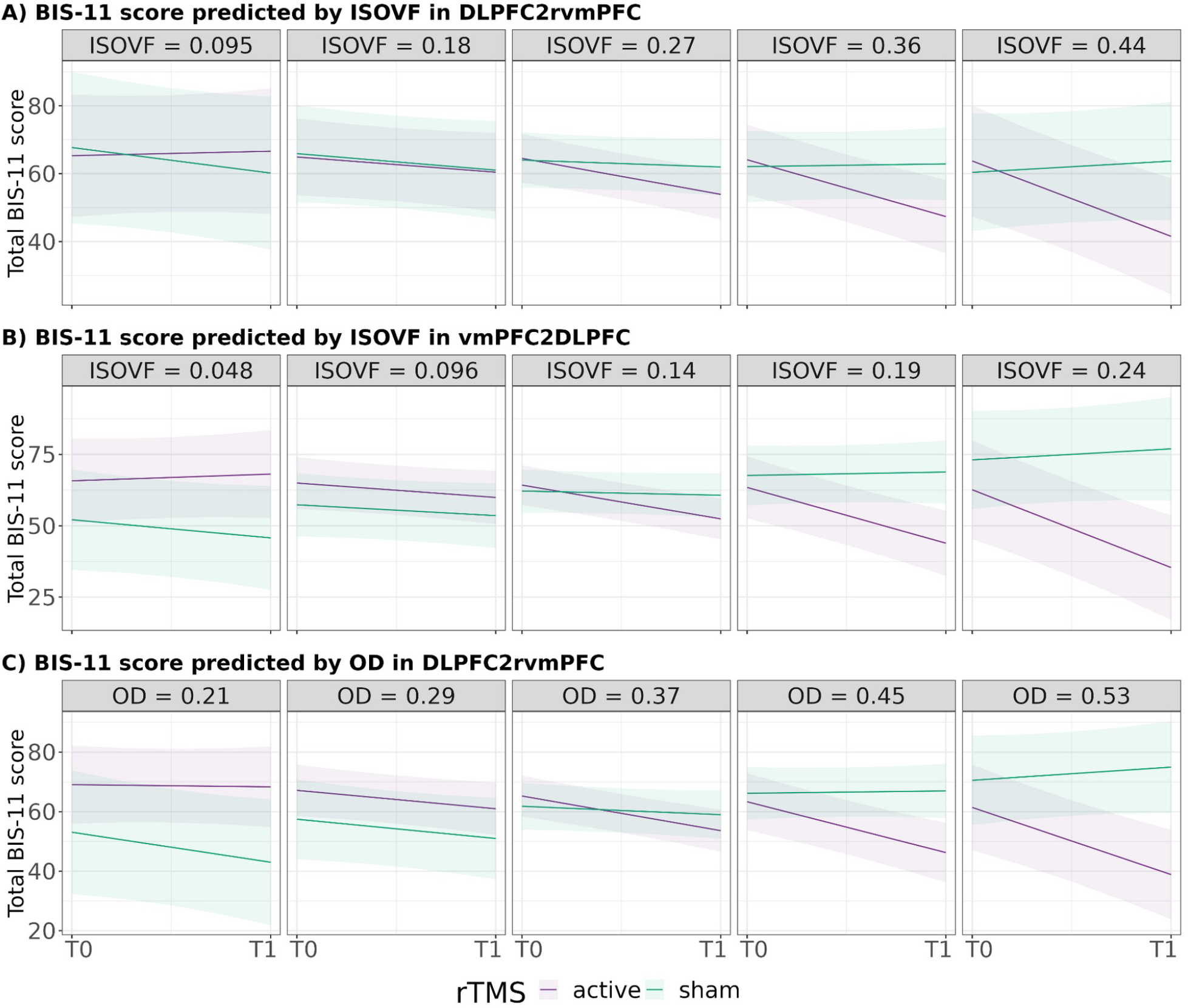
Prediction of total BIS-11 scores by the effect of rTMS on ISOVF/OD from frontostriatal circuits’ WM tracts. A) prediction of total BIS-11 scores from different ISOVF values in WM connecting left rostral middle frontal cortex and right medial orbitofrontal cortex (DLPFC2rvmPFC), B) prediction of total BIS-11 scores from different ISOVF values in WM connecting left right medial orbitofrontal cortex and left left rostral middle frontal cortex (vmPFC2DLPFC), C) prediction of total BIS-11 scores from different OD values in DLPFC2rvmPFC. BIS-11 = Barratt Impulsivity Scale v. 11, ISOVF = CSF volume fraction, OD = orientation dispersion, T0 = baseline time-point, T1 = after two weeks of rTMS protocol.

We found that BIS11 responsiveness seemed to be predicted by a high baseline ISOVF or free-water in WM connecting DLPFC with left (p=0.015, FDR=0.135, d=1.031) and right vmPFC (p=0.038, FDR=0.171, d=0.925) (**Table 3** and **Figure 3B**). None of these results regarding BIS11 survived multiple comparisons.

Finally, we only found that baseline OD from WM connecting left DLPFC and right vmPFC, when high, predicted greater decrease in BIS11 (p=0.020, FDR=0.180, d=0.827) (**Table 3** and **Figure 3C**), without surviving multiple comparisons. These results suggest that low baseline axon and dendrite density of subcortical tracts predict changes in craving secondary to rTMS, with high effect sizes.

## Discussion

Here, we investigated the effects of a 2-week treatment with rTMS over the left dorsolateral prefrontal cortex (lDLPFC) on fronto-striato-thalamic white matter (WM) microstructure in cocaine use disorder patients (CUD), using multishell diffusion-weighted imaging and NODDI measures. We further evaluated the correlation of clinical changes in craving and impulsivity with baseline WM microstructure. Our results suggest that rTMS on lDLPFC induces WM microstructural changes (density of axons and dendrites, and axon organization) in the lDLPFC-vmPFC and lDLPFC-left caudate nucleus connections after 2 weeks of treatment. We also found that baseline WM microstructure predicted clinical response to rTMS treatment. Low density of axons and dendrites in left caudate-substantia nigra, left caudate-left pallidum, left thalamus-substantia nigra connections predicted greater craving reductions. While we found more significant results, none survived multiple comparisons correction. We still discuss these results due to their effect size and possible significance in rTMS.

Repeated TMS showed an effect in WM tracts after 2 weeks, evidenced with an increase in ICVF (axon/dendrite density) and OD (axon orientation), but not ISOVF (free-water), in WM tracts connecting the lDLPFC to the ventromedial PFC (vmPFC) and to the left caudate nucleus in the Active group. We decided to discuss the ICVF result as it was significant at the individual level, with a multiple comparisons significance of 0.058, and a moderate to low effect size. These results may suggest a recovery of the integrity of prefrontal connections neurites with subcortical basal ganglia due to active rTMS as early as 2 weeks. The main mechanism of action proposed for rTMS in SUDs treatment is that it activates dopamine function in the meso-cortico-limbic system and an increase of extracellular glutamate and dopamine concentrations [10]. These changes could trigger changes in dendritic spine density, neurotransmitter receptor expression, neuronal degeneration, and microglial activation which could explain the results in basal ganglia and PFC [8,52,53]. For example, rTMS may affect calcium and neurotransmission levels. Further, Glutamate, GABA and some monoamines like dopamine (DA) have been reported to increase after PFC rTMS, leading to the dendritic spine enlargements through a signaling cascade [8,52]. DA is a critical neurotransmitter involved in synaptic plasticity in corticostriatal networks and in forming cue-induced craving, therefore DA stimulation-induced release over the striatum has been proposed as the mechanism underlying the effects of rTMS in substance use disorder patients [10,52].

A previous study has found that rTMS can reach deep subcortical structures such as the amygdala through stimulation of cortical regions by cortico-subcortical structural connections [54]. Although the strongest stimulation effects remain at the scalp level, and rapidly attenuates as it crosses to brain tissue, the activations spread to other sites via axonal connections and the releasing of neurotransmitters, leading to deep structural and functional network changes [10,55]. Our electric-field simulations showed that active rTMS in patients with CUD may have reached deep structures reflected by changes in NODDI metrics. Although we found these changes in WM tracts, they did not correlate to change in craving or impulsivity.

Baseline ICVF of WM tracts connecting left caudate nucleus-substantia nigra and left pallidum, and those connecting left thalamus-substantia nigra and left pallidum to frontal and subcortical regions predicted a reduction in craving VAS after the rTMS treatment. These results survived multiple comparisons and were the most robust. This was not the case for ICVF from WM tracts connecting cortical with subcortical regions. On the contrary, ISOVF predicted impulsivity responsiveness in left DLPFC connections to vmPFC and their connection of this to the right DLPFC. Similarly, OD predicted BIS11 responsiveness in one WM-ROI. However, these results did not survive multiple comparisons. We suggest that the initial state of WM integrity or the initial topology configuration are important for the rTMS outcome, or at least those patients had the highest changes after the therapy which may explain the higher effects. This may serve as an initial indication to select CUD patients that may more effectively benefit from this therapy. Prior studies have reported the association between subjective craving scores or impulsivity with diffusion metrics [56,57], with GM volumes [58,59], and with functional connectivity [60]. In which, they have suggested a detrimental effect of cocaine use on the brain that could indicate the status of craving and impulsivity, supporting the idea of WM-GM integrity as indicator of CUD severity and the guidance to achieving effective recovery.

Previous studies have found associations between repetitive stimulation over the DLPFC as well as the mPFC with improvements in executive functions, reward functions, as well as reduction of craving/drug consumption in patients with CUD [61,62]. The prediction of craving outcome based on the state of microstructure prior to stimulations could suggest that greater neurite integrity will result in better rTMS clinical outcomes, which is in line with previous studies that have found associations between WM-GM baseline integrity with better longitudinal clinical outcomes that can informs if patients will relapse or remain in drug abstinent [63]. This idea may be supported by the low effect size we found for NODDI changes, but high for NODDI as predictor. This may imply that irrespective of the induced changes, the initial integrity state has the highest impact on how rTMS modulates brain circuits and on the possible success of clinical outcome.

One of the main limitations of the present work concerns the longitudinal course, which is restricted to a single time point, only after 2 weeks. The results of which could be statistically confirmed with continued use of rTMS in the months of patient follow-up, as well as being extended to other regions. Even so, finding significant changes at 2 weeks suggests rTMS affects WM structure from the first day of stimulation. According to previous clinical disorders studies, ICVF is decreased with axonal degeneration, axonal loss and demyelination processes while OD decreasing reflects selective loss of crossing fibers, axonal regrowth and reduced axonal dispersion [12]. However, other researchers have found apparently contradictory results in terms of NODDI values directionality and the biological interpretation of it [12,16]. In this context, validation of NODDI values in terms of biological meaning would help to elucidate the current results. Discussion regarding the clinical sample variability and composition here used can be found in Garza-Villarreal et al. [21].

## Conclusion

In conclusion, the here shown reversive neurite microstructure state due to rTMS in connections between the prefrontal cortex and striatum, indicates a plasticity-induced recovery of the integrity of prefrontal connections by active stimulation. The prediction of the craving state based on baseline NODDI values before the onset of rTMS suggests that rTMS efficacy depends on the consumption-related integrity state of the underlying microstructure. Our results highlight rTMS as a potential therapeutic tool in the treatment of CUD, due to its ability to modulate altered brain microstructure by cocaine use, as well as the relevance of NODDI for tracing drug/treatment responsiveness.

## Data Availability

The MRI dataset can be found in https://openneuro.org/datasets/ds003037/. Clinical and cognitive measures are available in Zenodo https://doi.org/10.5281/zenodo.7126853.

## Supporting information

Supplementary rTMS method

## Data Availability

The MRI dataset can be found in https://openneuro.org/datasets/ds003037/

## Code Availability

For the code analysis presented here, please check: https://jalilrt.github.io/NODDI-changes-by_rTMS-in_CUD/

## Author contributions

Jalil Rasgado-Toledo: Formal analysis, Software, Methodology Visualization, Writing - original draft Victor Issa-Garcia: Formal analysis, Software, Methodology, Writing - original draft

Ruth Alcalá-Lozano: Funding acquisition, Investigation, Supervision, Project administration, Resources, Writing - review & editing.

Eduardo A. Garza-Villarreal: Conceptualization, Funding acquisition, Investigation, Supervision, Resources, Writing - review & editing.

Gabriel González-Escamilla: Conceptualization, Methodology, Investigation, Supervision, Writing - review & editing.

## Acknowledgements

The study was supported by public funds CONACYT FOSISS No. 0260971, CONACYT No. 253072 and PAPIIT-UNAM IA202120. Victor Issa-Garcia and Eduardo A. Garza-Villarreal would like to thank Dirección General de Calidad y Educación en Salud, Secretaría de Salud, México for the scholarship support provided to Victor. We also thank the Laboratorio Nacional de Visualización Científica Avanzada (LAVIS) for the use of their computer cluster and the Laboratorio Nacional de Imagenología por Resonancia Magnética (LANIREM). Gabriel Gonzalez-Escamilla is supported by a grant from the German Research Foundation (DFG; CRC-TR-128). Jalil Rasgado Toledo is a doctoral student from the Programa de Doctorado en Ciencias Biomédicas, Universidad Nacional Autónoma de México (UNAM) and received fellowship 858667 from CONACYT.

