## Supplementary tables and figures for "Cortical and subcortical connections change after repetitive transcranial magnetic stimulation therapy in cocaine use disorder and predict clinical outcome"

**Supplementary table 1.** Coefficients of linear mixed effects models’ interaction showing the effect of rTMS on neurite density (ICVF) from frontostriatal circuits’ white matter (WM) tracts.

| **weeks:rTMS[active]** | | | | | |
| --- | --- | --- | --- | --- | --- |
| **WM ROI** | **Estimate** | **Std. error** | **df** | **p-value** | **FDR** |
| Caud2Medulla | 0.006 | 0.005 | 41.65 | 0.288 | 0.495 |
| Caud2Palli | 0.003 | 0.004 | 41.47 | 0.495 | 0.636 |
| DLPFC2Caud | 0.010 | 0.006 | 42.69 | 0.105 | 0.315 |
| DLPFC2rvmPFC | 0.034 | 0.012 | 42.36 | 0.009* | 0.058 |
| DLPFC2Thal | 0.007 | 0.005 | 42.57 | 0.196 | 0.441 |
| rAngG2rDLPFC | -0.001 | 0.004 | 42.25 | 0.788 | 0.788 |
| Thal2Medulla | 0.007 | 0.007 | 41.61 | 0.330 | 0.495 |
| Thal2Palli | 0.002 | 0.005 | 41.39 | 0.740 | 0.788 |
| vmPFC2DLPFC | 0.038 | 0.015 | 42.31 | 0.013* | 0.058 |

Models: ${ICVF}_{{ROI}_{i}}=\beta_{0}+\beta_{1}\cdot weeks+\beta_{2}\cdot rTMS\left[ active \right]+\beta_{3}\cdot age+\beta_{4}\cdot educationyears+\beta_{5}\cdot weeks\cdot$ $rTMS\left[ active \right]$
Caud2Medulla: WM connecting left caudate nucleus and axial section through medulla, Caud2Palli: WM connecting left caudate nucleus and left pallidum, DLPFC2Caud: WM connecting left rostral middle frontal cortex (RMFC) and left caudate nucleus, DLPFC2rvmPFC: WM connecting left RMFC and right medial orbitofrontal cortex (MOC), DLPFC2Thal: WM connecting left RMFC and left thalamus, rAngG2rDLPFC: WM connecting right inferior parietal cortex and right RMFC, Thal2Medulla: WM connecting left thalamus and axial section through medulla, Thal2Palli: WM connecting left thalamus and left pallidum, vmPFC2DLPFC: WM connecting left MOC and left RMFC.

**Supplementary table 2.** Coefficients of linear mixed effects models’ interaction showing the effect of rTMS on CSF volume fraction (ISOVF) from frontostriatal circuits’ WM tracts.

| **weeks:rTMS[active]** | | | | | |
| --- | --- | --- | --- | --- | --- |
| **WM ROI** | **Estimate** | **Std. error** | **df** | **p-value** | **FDR** |
| Caud2Medulla | 0.001 | 0.005 | 41.76 | 0.792 | 0.792 |
| Caud2Palli | 0.002 | 0.003 | 41.46 | 0.496 | 0.638 |
| DLPFC2Caud | 0.002 | 0.003 | 41.62 | 0.489 | 0.638 |
| DLPFC2rvmPFC | 0.004 | 0.005 | 41.65 | 0.416 | 0.638 |
| DLPFC2Thal | 0.003 | 0.003 | 41.56 | 0.288 | 0.638 |
| rAngG2rDLPFC | -0.004 | 0.002 | 41.82 | 0.092 | 0.638 |
| Thal2Medulla | 0.003 | 0.006 | 42.03 | 0.612 | 0.689 |
| Thal2Palli | 0.004 | 0.005 | 41.74 | 0.348 | 0.638 |
| vmPFC2DLPFC | 0.008 | 0.005 | 42.13 | 0.155 | 0.638 |

Models: ${ISOVF}_{{ROI}_{i}}=\beta_{0}+\beta_{1}\cdot weeks+\beta_{2}\cdot rTMS\left[ active \right]+\beta_{3}\cdot age+\beta_{4}\cdot educationyears+\beta_{5}\cdot weeks\cdot$ $rTMS\left[ active \right]$Caud2Medulla: WM connecting left caudate nucleus and axial section through medulla, Caud2Palli: WM connecting left caudate nucleus and left pallidum, DLPFC2Caud: WM connecting left rostral middle frontal cortex (RMFC) and left caudate nucleus, DLPFC2rvmPFC: WM connecting left RMFC and right medial orbitofrontal cortex (MOC), DLPFC2Thal: WM connecting left RMFC and left thalamus, rAngG2rDLPFC: WM connecting right inferior parietal cortex and right RMFC, Thal2Medulla: WM connecting left thalamus and axial section through medulla, Thal2Palli: WM connecting left thalamus and left pallidum, vmPFC2DLPFC: WM connecting left MOC and left RMFC.

**Supplementary table 3.** Coefficients of linear mixed effects models’ interaction showing the effect of rTMS on orientation dispersion (OD) from frontostriatal circuits’ WM tracts.

| **weeks:rTMS[active]** | | | | | |
| --- | --- | --- | --- | --- | --- |
| **WM ROI** | **Estimate** | **Std. error** | **df** | **p-value** | **FDR** |
| Caud2Medulla | -0.0003 | 0.004 | 41.94 | 0.949 | 0.949 |
| Caud2Palli | 0.002 | 0.002 | 41.33 | 0.433 | 0.557 |
| DLPFC2Caud | 0.009 | 0.004 | 40.65 | 0.021* | 0.094 |
| DLPFC2rvmPFC | 0.013 | 0.007 | 42.18 | 0.076 | 0.171 |
| DLPFC2Thal | 0.006 | 0.003 | 40.68 | 0.045* | 0.135 |
| rAngG2rDLPFC | 0.003 | 0.003 | 42.34 | 0.204 | 0.367 |
| Thal2Medulla | -0.001 | 0.005 | 42.54 | 0.809 | 0.910 |
| Thal2Palli | 0.003 | 0.003 | 41.76 | 0.380 | 0.557 |
| vmPFC2DLPFC | 0.027 | 0.010 | 41.91 | 0.012* | 0.094 |

Models: ${OD}_{{ROI}_{i}}=\beta_{0}+\beta_{1}\cdot weeks+\beta_{2}\cdot rTMS\left[ active \right]+\beta_{3}\cdot age+\beta_{4}\cdot educationyears+\beta_{5}\cdot weeks\cdot$ $rTMS\left[ active \right]$
Caud2Medulla: WM connecting left caudate nucleus and axial section through medulla, Caud2Palli: WM connecting left caudate nucleus and left pallidum, DLPFC2Caud: WM connecting left rostral middle frontal cortex (RMFC) and left caudate nucleus, DLPFC2rvmPFC: WM connecting left RMFC and right medial orbitofrontal cortex (MOC), DLPFC2Thal: WM connecting left RMFC and left thalamus, rAngG2rDLPFC: WM connecting right inferior parietal cortex and right RMFC, Thal2Medulla: WM connecting left thalamus and axial section through medulla, Thal2Palli: WM connecting left thalamus and left pallidum, vmPFC2DLPFC: WM connecting left MOC and left RMFC.

**Supplementary table 4.** Coefficients of linear mixed effects models’ 3-way interaction predicting decrease in craving VAS using baseline neurite density (ICVF) from frontostriatal circuits’ WM tracts.

| **weeks:ICVF:rTMS[active]** | | | | | |
| --- | --- | --- | --- | --- | --- |
| **WM ROI** | **Estimate** | **Std. error** | **df** | **p-value** | **FDR** |
| Caud2Medulla | 22.61 | 7.93 | 40.12 | 0.007* | 0.021* |
| Caud2Palli | 21.13 | 6.95 | 40.17 | 0.004* | 0.018* |
| DLPFC2Caud | 1.28 | 6.33 | 38.44 | 0.841 | 0.841 |
| DLPFC2rvmPFC | -1.36 | 3.40 | 40.15 | 0.691 | 0.777 |
| DLPFC2Thal | 10.00 | 8.14 | 38.97 | 0.226 | 0.407 |
| rAngG2rDLPFC | 5.22 | 9.14 | 38.85 | 0.571 | 0.777 |
| Thal2Medulla | 16.30 | 6.17 | 40.07 | 0.012* | 0.027* |
| Thal2Palli | 21.92 | 6.61 | 40.13 | 0.002* | 0.018* |
| vmPFC2DLPFC | -1.35 | 3.16 | 40.22 | 0.670 | 0.777 |

Models: $VAS=\beta_{0}+\beta_{1}\cdot weeks+\beta_{2}\cdot{ICVF}_{{ROI}_{i}}+\beta_{3}\cdot rTMS\left[ active \right]+\beta_{4}\cdot age+\beta_{5}\cdot educationyears+\beta_{6}\cdot$
$weeks\cdot{ICVF}_{{ROI}_{i}}+\beta_{7}\cdot weeks\cdot rTMS\left[ active \right]+\beta_{8}\cdot{ICVF}_{{ROI}_{i}}\cdot rTMS\left[ active \right]+\beta_{9}\cdot{ICVF}_{{ROI}_{i}}\cdot age+$
$\beta_{10}\cdot{ICVF}_{{ROI}_{i}}\cdot educationyears+\beta_{11}\cdot weeks\cdot{ICVF}_{{ROI}_{i}}\cdot rTMS\left[ active \right]$
Caud2Medulla: WM connecting left caudate nucleus and axial section through medulla, Caud2Palli: WM connecting left caudate nucleus and left pallidum, DLPFC2Caud: WM connecting left rostral middle frontal cortex (RMFC) and left caudate nucleus, DLPFC2rvmPFC: WM connecting left RMFC and right medial orbitofrontal cortex (MOC), DLPFC2Thal: WM connecting left RMFC and left thalamus, rAngG2rDLPFC: WM connecting right inferior parietal cortex and right RMFC, Thal2Medulla: WM connecting left thalamus and axial section through medulla, Thal2Palli: WM connecting left thalamus and left pallidum, vmPFC2DLPFC: WM connecting left MOC and left RMFC.

**Supplementary table 5.** Coefficients of linear mixed effects models’ 3-way interaction predicting decrease in craving CCQ-now by using baseline neurite density (ICVF) from frontostriatal circuits’ WM tracts.

| **weeks:ICVF:rTMS[active]** | | | | | |
| --- | --- | --- | --- | --- | --- |
| **WM ROI** | **Estimate** | **Std. error** | **df** | **p-value** | **FDR** |
| Caud2Medulla | 165.11 | 129.68 | 39.60 | 0.210 | 0.630 |
| Caud2Palli | 129.75 | 118.63 | 39.42 | 0.281 | 0.632 |
| DLPFC2Caud | -2.13 | 108.27 | 38.57 | 0.984 | 0.984 |
| DLPFC2rvmPFC | 1.84 | 52.20 | 39.94 | 0.972 | 0.984 |
| DLPFC2Thal | 73.69 | 137.02 | 38.39 | 0.594 | 0.973 |
| rAngG2rDLPFC | 61.23 | 144.70 | 38.60 | 0.675 | 0.973 |
| Thal2Medulla | 173.07 | 99.50 | 39.49 | 0.090 | 0.630 |
| Thal2Palli | 154.50 | 113.10 | 39.98 | 0.180 | 0.630 |
| vmPFC2DLPFC | -15.80 | 50.69 | 40.57 | 0.757 | 0.973 |

Models: $CCQnow=\beta_{0}+\beta_{1}\cdot weeks+\beta_{2}\cdot{ICVF}_{{ROI}_{i}}+\beta_{3}\cdot rTMS\left[ active \right]+\beta_{4}\cdot age+\beta_{5}\cdot educationyears+$
$\beta_{6}\cdot weeks\cdot{ICVF}_{{ROI}_{i}}+\beta_{7}\cdot weeks\cdot rTMS\left[ active \right]+\beta_{8}\cdot{ICVF}_{{ROI}_{i}}\cdot rTMS\left[ active \right]+\beta_{9}\cdot{ICVF}_{{ROI}_{i}}\cdot age+$
$\beta_{10}\cdot{ICVF}_{{ROI}_{i}}\cdot educationyears+\beta_{11}\cdot weeks\cdot{ICVF}_{{ROI}_{i}}\cdot rTMS\left[ active \right]$
Caud2Medulla: WM connecting left caudate nucleus and axial section through medulla, Caud2Palli: WM connecting left caudate nucleus and left pallidum, DLPFC2Caud: WM connecting left rostral middle frontal cortex (RMFC) and left caudate nucleus, DLPFC2rvmPFC: WM connecting left RMFC and right medial orbitofrontal cortex (MOC), DLPFC2Thal: WM connecting left RMFC and left thalamus, rAngG2rDLPFC: WM connecting right inferior parietal cortex and right RMFC, Thal2Medulla: WM connecting left thalamus and axial section through medulla, Thal2Palli: WM connecting left thalamus and left pallidum, vmPFC2DLPFC: WM connecting left MOC and left RMFC.

**Supplementary table 6.** Coefficients of linear mixed effects models’ 3-way interaction predicting decrease in total BIS11 by using baseline neurite density (ICVF) from frontostriatal circuits’ WM tracts. Models:

| **weeks:ICVF:rTMS[active]** | | | | | |
| --- | --- | --- | --- | --- | --- |
| **WM ROI** | **Estimate** | **Std. error** | **df** | **p-value** | **FDR** |
| Caud2Medulla | 30.38 | 39.46 | 40.41 | 0.446 | 0.647 |
| Caud2Palli | 23.98 | 35.50 | 40.24 | 0.503 | 0.647 |
| DLPFC2Caud | -33.83 | 32.05 | 39.12 | 0.298 | 0.536 |
| DLPFC2rvmPFC | -24.06 | 15.62 | 40.82 | 0.131 | 0.480 |
| DLPFC2Thal | -13.12 | 40.86 | 39.38 | 0.750 | 0.753 |
| rAngG2rDLPFC | 13.71 | 43.27 | 39.74 | 0.753 | 0.753 |
| Thal2Medulla | 35.96 | 30.23 | 40.28 | 0.241 | 0.536 |
| Thal2Palli | 48.04 | 33.59 | 40.52 | 0.160 | 0.480 |
| vmPFC2DLPFC | -27.13 | 15.19 | 40.53 | 0.081 | 0.480 |

$BIS11=\beta_{0}+\beta_{1}\cdot weeks+\beta_{2}\cdot{ICVF}_{{ROI}_{i}}+\beta_{3}\cdot rTMS\left[ active \right]+\beta_{4}\cdot age+\beta_{5}\cdot educationyears+\beta_{6}\cdot$
$weeks\cdot{ICVF}_{{ROI}_{i}}+\beta_{7}\cdot weeks\cdot rTMS\left[ active \right]+\beta_{8}\cdot{ICVF}_{{ROI}_{i}}\cdot rTMS\left[ active \right]+\beta_{9}\cdot{ICVF}_{{ROI}_{i}}\cdot age+$
$\beta_{10}\cdot{ICVF}_{{ROI}_{i}}\cdot educationyears+\beta_{11}\cdot weeks\cdot{ICVF}_{{ROI}_{i}}\cdot rTMS\left[ active \right]$
Caud2Medulla: WM connecting left caudate nucleus and axial section through medulla, Caud2Palli: WM connecting left caudate nucleus and left pallidum, DLPFC2Caud: WM connecting left rostral middle frontal cortex (RMFC) and left caudate nucleus, DLPFC2rvmPFC: WM connecting left RMFC and right medial orbitofrontal cortex (MOC), DLPFC2Thal: WM connecting left RMFC and left thalamus, rAngG2rDLPFC: WM connecting right inferior parietal cortex and right RMFC, Thal2Medulla: WM connecting left thalamus and axial section through medulla, Thal2Palli: WM connecting left thalamus and left pallidum, vmPFC2DLPFC: WM connecting left MOC and left RMFC.

**Supplementary table 7.** Coefficients of linear mixed effects models’ 3-way interaction predicting decrease in craving VAS using baseline CSF volume fraction (ISOVF) from frontostriatal circuits’ WM tracts.

| **weeks:ISOVF:rTMS[active]** | | | | | |
| --- | --- | --- | --- | --- | --- |
| **WM ROI** | **Estimate** | **Std. error** | **df** | **p-value** | **FDR** |
| Caud2Medulla | 3.35 | 6.28 | 39.23 | 0.597 | 0.931 |
| Caud2Palli | -2.31 | 6.72 | 39.08 | 0.732 | 0.931 |
| DLPFC2Caud | 5.40 | 9.41 | 40.19 | 0.569 | 0.931 |
| DLPFC2rvmPFC | -2.31 | 5.24 | 38.52 | 0.662 | 0.931 |
| DLPFC2Thal | 6.40 | 8.86 | 39.04 | 0.474 | 0.931 |
| rAngG2rDLPFC | 16.80 | 12.37 | 39.35 | 0.182 | 0.931 |
| Thal2Medulla | 5.75 | 5.52 | 38.91 | 0.304 | 0.931 |
| Thal2Palli | 1.26 | 5.76 | 38.62 | 0.828 | 0.931 |
| vmPFC2DLPFC | 0.54 | 9.18 | 39.70 | 0.953 | 0.953 |

Models: $VAS=\beta_{0}+\beta_{1}\cdot weeks+\beta_{2}\cdot{ISOVF}_{{ROI}_{i}}+\beta_{3}\cdot rTMS\left[ active \right]+\beta_{4}\cdot age+\beta_{5}\cdot educationyears+\beta_{6}\cdot$
$weeks\cdot{ISOVF}_{{ROI}_{i}}+\beta_{7}\cdot weeks\cdot rTMS\left[ active \right]+\beta_{8}\cdot{ISOVF}_{{ROI}_{i}}\cdot rTMS\left[ active \right]+\beta_{9}\cdot{ISOVF}_{{ROI}_{i}}\cdot age+$
$\beta_{10}\cdot{ISOVF}_{{ROI}_{i}}\cdot educationyears+\beta_{11}\cdot weeks\cdot{ISOVF}_{{ROI}_{i}}\cdot rTMS\left[ active \right]$
Caud2Medulla: WM connecting left caudate nucleus and axial section through medulla, Caud2Palli: WM connecting left caudate nucleus and left pallidum, DLPFC2Caud: WM connecting left rostral middle frontal cortex (RMFC) and left caudate nucleus, DLPFC2rvmPFC: WM connecting left RMFC and right medial orbitofrontal cortex (MOC), DLPFC2Thal: WM connecting left RMFC and left thalamus, rAngG2rDLPFC: WM connecting right inferior parietal cortex and right RMFC, Thal2Medulla: WM connecting left thalamus and axial section through medulla, Thal2Palli: WM connecting left thalamus and left pallidum, vmPFC2DLPFC: WM connecting left MOC and left RMFC.

**Supplementary table 8.** Coefficients of linear mixed effects models’ 3-way interaction predicting decrease in craving CCQ-now by using baseline CSF volume fraction (ISOVF) from frontostriatal circuits’ WM tracts.

| **weeks:ISOVF:rTMS[active]** | | | | | |
| --- | --- | --- | --- | --- | --- |
| **WM ROI** | **Estimate** | **Std. error** | **df** | **p-value** | **FDR** |
| Caud2Medulla | -80.27 | 94.20 | 38.48 | 0.399 | 0.736 |
| Caud2Palli | -105.69 | 99.70 | 38.64 | 0.296 | 0.736 |
| DLPFC2Caud | -117.91 | 141.26 | 39.65 | 0.409 | 0.736 |
| DLPFC2rvmPFC | -161.10 | 79.34 | 38.92 | 0.049* | 0.441 |
| DLPFC2Thal | -44.27 | 133.19 | 38.59 | 0.741 | 0.807 |
| rAngG2rDLPFC | -108.75 | 182.55 | 38.53 | 0.555 | 0.807 |
| Thal2Medulla | -29.53 | 83.28 | 38.00 | 0.725 | 0.807 |
| Thal2Palli | -20.97 | 85.18 | 38.13 | 0.807 | 0.807 |
| vmPFC2DLPFC | -209.04 | 145.77 | 40.52 | 0.159 | 0.716 |

Models: $CCQnow=\beta_{0}+\beta_{1}\cdot weeks+\beta_{2}\cdot{ISOVF}_{{ROI}_{i}}+\beta_{3}\cdot rTMS\left[ active \right]+\beta_{4}\cdot age+\beta_{5}\cdot educationyears+$
$\beta_{6}\cdot weeks\cdot{ISOVF}_{{ROI}_{i}}+\beta_{7}\cdot weeks\cdot rTMS\left[ active \right]+\beta_{8}\cdot{ISOVF}_{{ROI}_{i}}\cdot rTMS\left[ active \right]+\beta_{9}\cdot{ISOVF}_{{ROI}_{i}}\cdot$
$age+\beta_{10}\cdot{ISOVF}_{{ROI}_{i}}\cdot educationyears+\beta_{11}\cdot weeks\cdot{ISOVF}_{{ROI}_{i}}\cdot rTMS\left[ active \right]$
Caud2Medulla: WM connecting left caudate nucleus and axial section through medulla, Caud2Palli: WM connecting left caudate nucleus and left pallidum, DLPFC2Caud: WM connecting left rostral middle frontal cortex (RMFC) and left caudate nucleus, DLPFC2rvmPFC: WM connecting left RMFC and right medial orbitofrontal cortex (MOC), DLPFC2Thal: WM connecting left RMFC and left thalamus, rAngG2rDLPFC: WM connecting right inferior parietal cortex and right RMFC, Thal2Medulla: WM connecting left thalamus and axial section through medulla, Thal2Palli: WM connecting left thalamus and left pallidum, vmPFC2DLPFC: WM connecting left MOC and left RMFC.

**Supplementary table 9.** Coefficients of linear mixed effects models’ 3-way interaction predicting decrease in total BIS11 by using baseline CSF volume fraction (ISOVF) from frontostriatal circuits’ WM tracts.

| **weeks:ISOVF:rTMS[active]** | | | | | |
| --- | --- | --- | --- | --- | --- |
| **WM ROI** | **Estimate** | **Std. error** | **df** | **p-value** | **FDR** |
| Caud2Medulla | -28.17 | 27.97 | 39.59 | 0.320 | 0.588 |
| Caud2Palli | -27.50 | 30.29 | 39.52 | 0.369 | 0.588 |
| DLPFC2Caud | -35.01 | 42.73 | 40.20 | 0.417 | 0.588 |
| DLPFC2rvmPFC | -49.81 | 23.25 | 39.63 | 0.038* | 0.171 |
| DLPFC2Thal | -26.44 | 39.94 | 39.90 | 0.512 | 0.588 |
| rAngG2rDLPFC | -2.35 | 57.45 | 39.83 | 0.968 | 0.968 |
| Thal2Medulla | -16.03 | 24.87 | 39.67 | 0.523 | 0.588 |
| Thal2Palli | -22.45 | 25.53 | 39.59 | 0.384 | 0.588 |
| vmPFC2DLPFC | -103.78 | 40.96 | 40.51 | 0.015* | 0.135 |

Models: $BIS11=\beta_{0}+\beta_{1}\cdot weeks+\beta_{2}\cdot{ISOVF}_{{ROI}_{i}}+\beta_{3}\cdot rTMS\left[ active \right]+\beta_{4}\cdot age+\beta_{5}\cdot educationyears+\beta_{6}\cdot$
$weeks\cdot{ISOVF}_{{ROI}_{i}}+\beta_{7}\cdot weeks\cdot rTMS\left[ active \right]+\beta_{8}\cdot{ISOVF}_{{ROI}_{i}}\cdot rTMS\left[ active \right]+\beta_{9}\cdot{ISOVF}_{{ROI}_{i}}\cdot age+$
$\beta_{10}\cdot{ISOVF}_{{ROI}_{i}}\cdot educationyears+\beta_{11}\cdot weeks\cdot{ISOVF}_{{ROI}_{i}}\cdot rTMS\left[ active \right]$
Caud2Medulla: WM connecting left caudate nucleus and axial section through medulla, Caud2Palli: WM connecting left caudate nucleus and left pallidum, DLPFC2Caud: WM connecting left rostral middle frontal cortex (RMFC) and left caudate nucleus, DLPFC2rvmPFC: WM connecting left RMFC and right medial orbitofrontal cortex (MOC), DLPFC2Thal: WM connecting left RMFC and left thalamus, rAngG2rDLPFC: WM connecting right inferior parietal cortex and right RMFC, Thal2Medulla: WM connecting left thalamus and axial section through medulla, Thal2Palli: WM connecting left thalamus and left pallidum, vmPFC2DLPFC: WM connecting left MOC and left RMFC.

**Supplementary table 10.** Coefficients of linear mixed effects models’ 3-way interaction predicting decrease in craving VAS using baseline orientation dispersion (OD) from frontostriatal circuits’ WM tracts.

| **weeks:OD:rTMS[active]** | | | | | |
| --- | --- | --- | --- | --- | --- |
| **WM ROI** | **Estimate** | **Std. error** | **df** | **p-value** | **FDR** |
| Caud2Medulla | 14.66 | 10.48 | 40.20 | 0.170 | 0.788 |
| Caud2Palli | 15.42 | 15.63 | 41.63 | 0.330 | 0.788 |
| DLPFC2Caud | 3.26 | 11.65 | 41.81 | 0.781 | 0.836 |
| DLPFC2rvmPFC | -3.14 | 5.12 | 39.30 | 0.544 | 0.788 |
| DLPFC2Thal | 2.85 | 13.72 | 41.86 | 0.836 | 0.836 |
| rAngG2rDLPFC | 16.78 | 17.47 | 41.86 | 0.342 | 0.788 |
| Thal2Medulla | 5.08 | 7.95 | 38.96 | 0.527 | 0.788 |
| Thal2Palli | 7.55 | 12.36 | 39.83 | 0.545 | 0.788 |
| vmPFC2DLPFC | -2.32 | 4.55 | 40.70 | 0.613 | 0.788 |

Models: $VAS=\beta_{0}+\beta_{1}\cdot weeks+\beta_{2}\cdot{OD}_{{ROI}_{i}}+\beta_{3}\cdot rTMS\left[ active \right]+\beta_{4}\cdot age+\beta_{5}\cdot educationyears+\beta_{6}\cdot$
$weeks\cdot{OD}_{{ROI}_{i}}+\beta_{7}\cdot weeks\cdot rTMS\left[ active \right]+\beta_{8}\cdot{OD}_{{ROI}_{i}}\cdot rTMS\left[ active \right]+\beta_{9}\cdot{OD}_{{ROI}_{i}}\cdot age+\beta_{10}\cdot$
${OD}_{{ROI}_{i}}\cdot educationyears+\beta_{11}\cdot weeks\cdot{OD}_{{ROI}_{i}}\cdot rTMS\left[ active \right]$
Caud2Medulla: WM connecting left caudate nucleus and axial section through medulla, Caud2Palli: WM connecting left caudate nucleus and left pallidum, DLPFC2Caud: WM connecting left rostral middle frontal cortex (RMFC) and left caudate nucleus, DLPFC2rvmPFC: WM connecting left RMFC and right medial orbitofrontal cortex (MOC), DLPFC2Thal: WM connecting left RMFC and left thalamus, rAngG2rDLPFC: WM connecting right inferior parietal cortex and right RMFC, Thal2Medulla: WM connecting left thalamus and axial section through medulla, Thal2Palli: WM connecting left thalamus and left pallidum, vmPFC2DLPFC: WM connecting left MOC and left RMFC.

**Supplementary table 11.** Coefficients of linear mixed effects models’ 3-way interaction predicting decrease in craving CCQ-now by using baseline orientation dispersion (OD) from frontostriatal circuits’ WM tracts.

| **weeks:OD:rTMS[active]** | | | | | |
| --- | --- | --- | --- | --- | --- |
| **WM ROI** | **Estimate** | **Std. error** | **df** | **p-value** | **FDR** |
| Caud2Medulla | 6.35 | 165.47 | 38.93 | 0.970 | 0.970 |
| Caud2Palli | 59.74 | 243.19 | 40.27 | 0.807 | 0.970 |
| DLPFC2Caud | -96.62 | 185.70 | 42.00 | 0.606 | 0.970 |
| DLPFC2rvmPFC | -82.64 | 81.99 | 39.51 | 0.320 | 0.970 |
| DLPFC2Thal | -115.30 | 221.16 | 41.66 | 0.605 | 0.970 |
| rAngG2rDLPFC | -350.29 | 261.71 | 40.27 | 0.188 | 0.970 |
| Thal2Medulla | -15.25 | 122.58 | 37.77 | 0.902 | 0.970 |
| Thal2Palli | 104.11 | 190.20 | 38.45 | 0.587 | 0.970 |
| vmPFC2DLPFC | 29.48 | 68.87 | 40.37 | 0.671 | 0.970 |

Models: $CCQnow=\beta_{0}+\beta_{1}\cdot weeks+\beta_{2}\cdot{OD}_{{ROI}_{i}}+\beta_{3}\cdot rTMS\left[ active \right]+\beta_{4}\cdot age+\beta_{5}\cdot educationyears+\beta_{6}\cdot$
$weeks\cdot{OD}_{{ROI}_{i}}+\beta_{7}\cdot weeks\cdot rTMS\left[ active \right]+\beta_{8}\cdot{OD}_{{ROI}_{i}}\cdot rTMS\left[ active \right]+\beta_{9}\cdot{OD}_{{ROI}_{i}}\cdot age+\beta_{10}\cdot$
${OD}_{{ROI}_{i}}\cdot educationyears+\beta_{11}\cdot weeks\cdot{OD}_{{ROI}_{i}}\cdot rTMS\left[ active \right]$
Caud2Medulla: WM connecting left caudate nucleus and axial section through medulla, Caud2Palli: WM connecting left caudate nucleus and left pallidum, DLPFC2Caud: WM connecting left rostral middle frontal cortex (RMFC) and left caudate nucleus, DLPFC2rvmPFC: WM connecting left RMFC and right medial orbitofrontal cortex (MOC), DLPFC2Thal: WM connecting left RMFC and left thalamus, rAngG2rDLPFC: WM connecting right inferior parietal cortex and right RMFC, Thal2Medulla: WM connecting left thalamus and axial section through medulla, Thal2Palli: WM connecting left thalamus and left pallidum, vmPFC2DLPFC: WM connecting left MOC and left RMFC.

**Supplementary table 12.** Coefficients of linear mixed effects models’ 3-way interaction predicting decrease in total BIS11 by using baseline orientation dispersion (OD) from frontostriatal circuits’ WM tracts.

| **weeks:OD:rTMS[active]** | | | | | |
| --- | --- | --- | --- | --- | --- |
| **WM ROI** | **Estimate** | **Std. error** | **df** | **p-value** | **FDR** |
| Caud2Medulla | -7.73 | 48.65 | 39.86 | 0.875 | 0.933 |
| Caud2Palli | 43.38 | 72.61 | 40.73 | 0.554 | 0.831 |
| DLPFC2Caud | -100.55 | 56.26 | 41.87 | 0.081 | 0.234 |
| DLPFC2rvmPFC | -56.76 | 23.43 | 39.80 | 0.020* | 0.180 |
| DLPFC2Thal | -109.30 | 65.74 | 41.73 | 0.104 | 0.234 |
| rAngG2rDLPFC | -48.97 | 82.08 | 41.03 | 0.554 | 0.831 |
| Thal2Medulla | -14.57 | 35.77 | 39.53 | 0.686 | 0.882 |
| Thal2Palli | -4.72 | 55.77 | 40.22 | 0.933 | 0.933 |
| vmPFC2DLPFC | -38.26 | 21.50 | 40.93 | 0.083 | 0.234 |

Models: $BIS11=\beta_{0}+\beta_{1}\cdot weeks+\beta_{2}\cdot{OD}_{{ROI}_{i}}+\beta_{3}\cdot rTMS\left[ active \right]+\beta_{4}\cdot age+\beta_{5}\cdot educationyears+\beta_{6}\cdot$
$weeks\cdot{OD}_{{ROI}_{i}}+\beta_{7}\cdot weeks\cdot rTMS\left[ active \right]+\beta_{8}\cdot{OD}_{{ROI}_{i}}\cdot rTMS\left[ active \right]+\beta_{9}\cdot{OD}_{{ROI}_{i}}\cdot age+\beta_{10}\cdot$
${OD}_{{ROI}_{i}}\cdot educationyears+\beta_{11}\cdot weeks\cdot{OD}_{{ROI}_{i}}\cdot rTMS\left[ active \right]$
Caud2Medulla: WM connecting left caudate nucleus and axial section through medulla, Caud2Palli: WM connecting left caudate nucleus and left pallidum, DLPFC2Caud: WM connecting left rostral middle frontal cortex (RMFC) and left caudate nucleus, DLPFC2rvmPFC: WM connecting left RMFC and right medial orbitofrontal cortex (MOC), DLPFC2Thal: WM connecting left RMFC and left thalamus, rAngG2rDLPFC: WM connecting right inferior parietal cortex and right RMFC, Thal2Medulla: WM connecting left thalamus and axial section through medulla, Thal2Palli: WM connecting left thalamus and left pallidum, vmPFC2DLPFC: WM connecting left MOC and left RMFC.

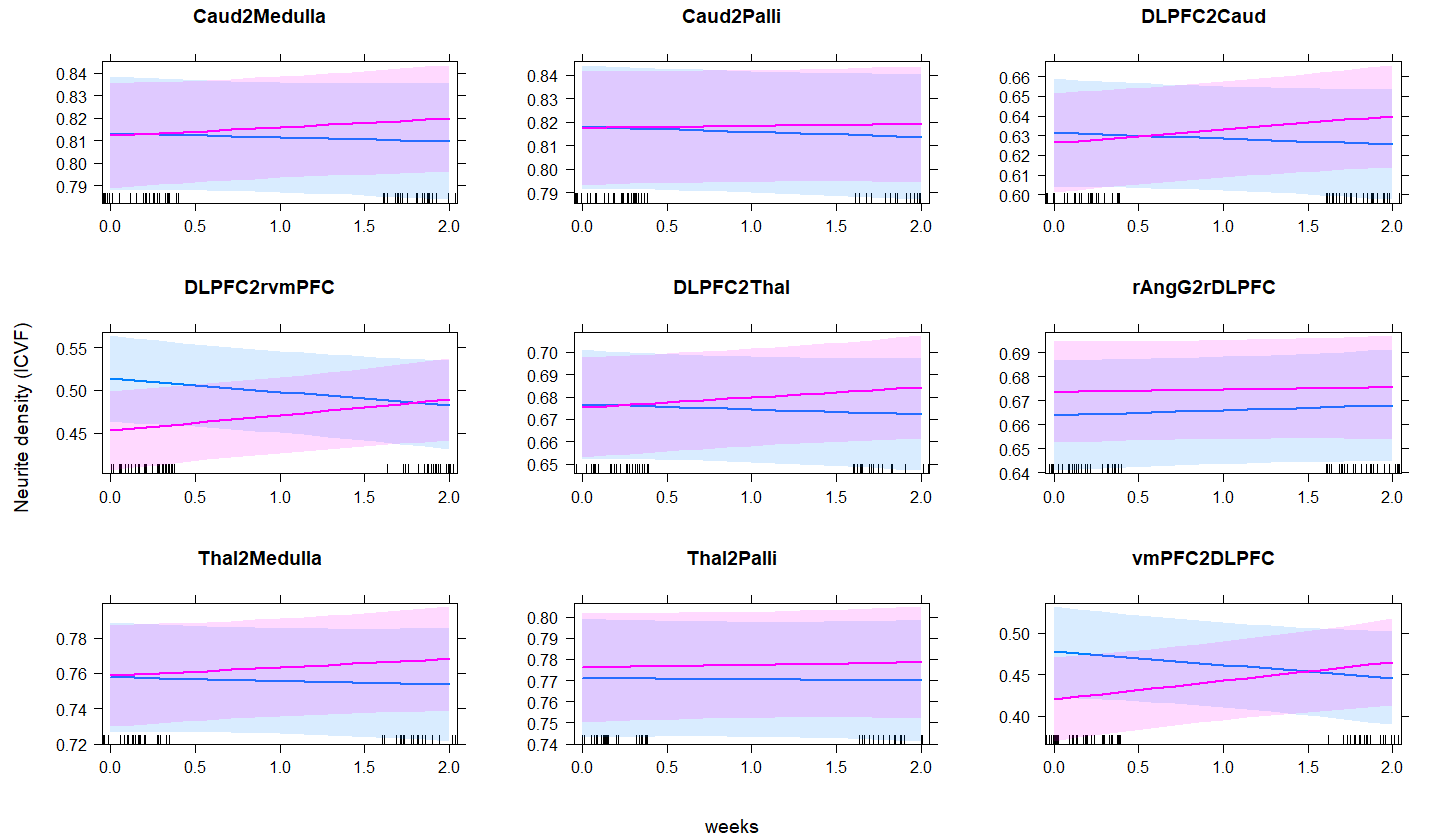

**Supplementary figure 1.** Effect of rTMS on neurite density (ICVF) from frontostriatal circuits’ WM tracts; blue = sham, pink = active

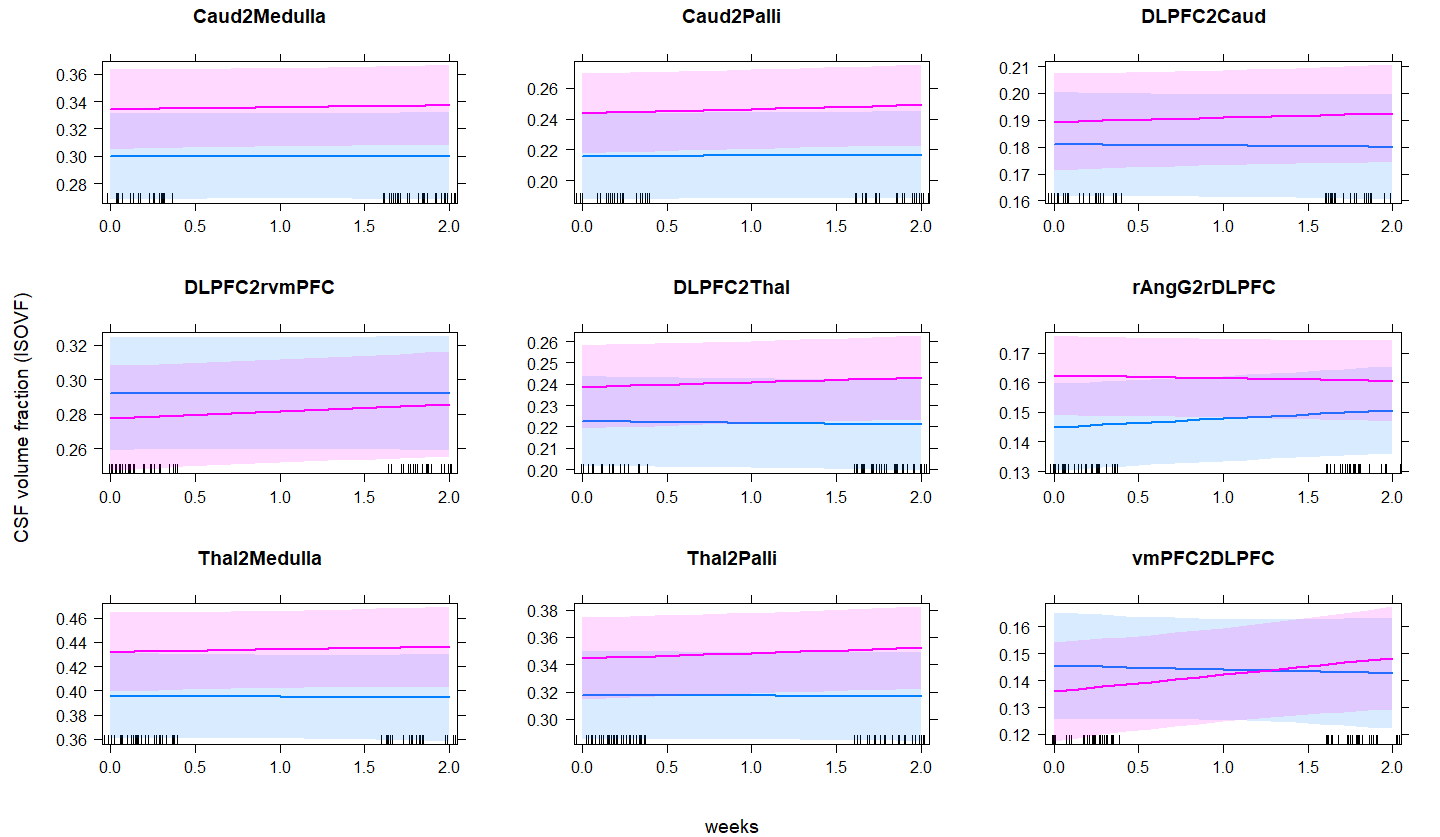

**Supplementary figure 2.** Effect of rTMS on CSF volume fraction (ISOVF) from frontostriatal circuits’ WM tracts; blue = sham, pink = active.

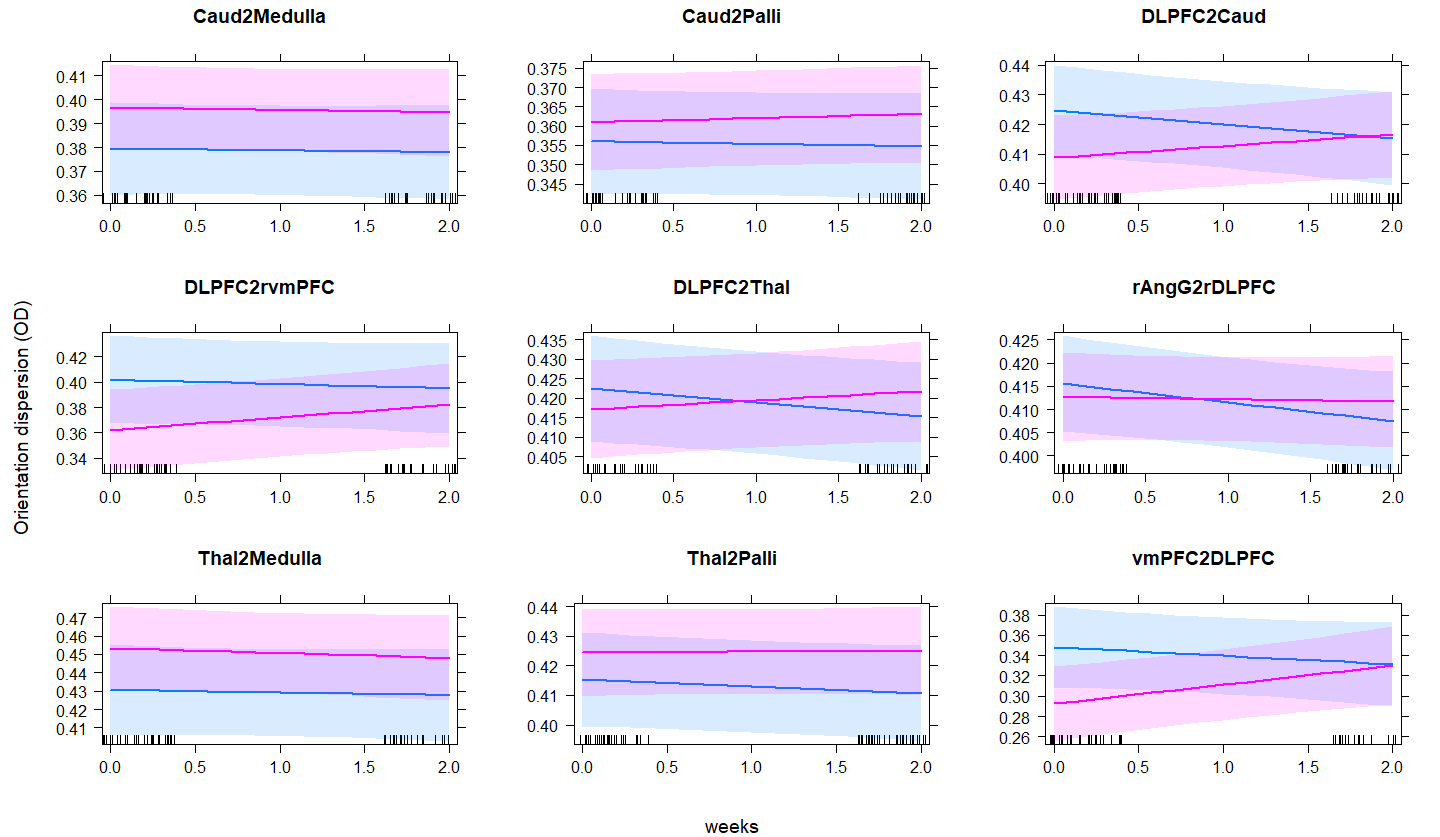

**Supplementary figure 3.** Effect of rTMS on orientation dispersion (OD) from frontostriatal circuits’ WM tracts; blue = sham, pink = active.

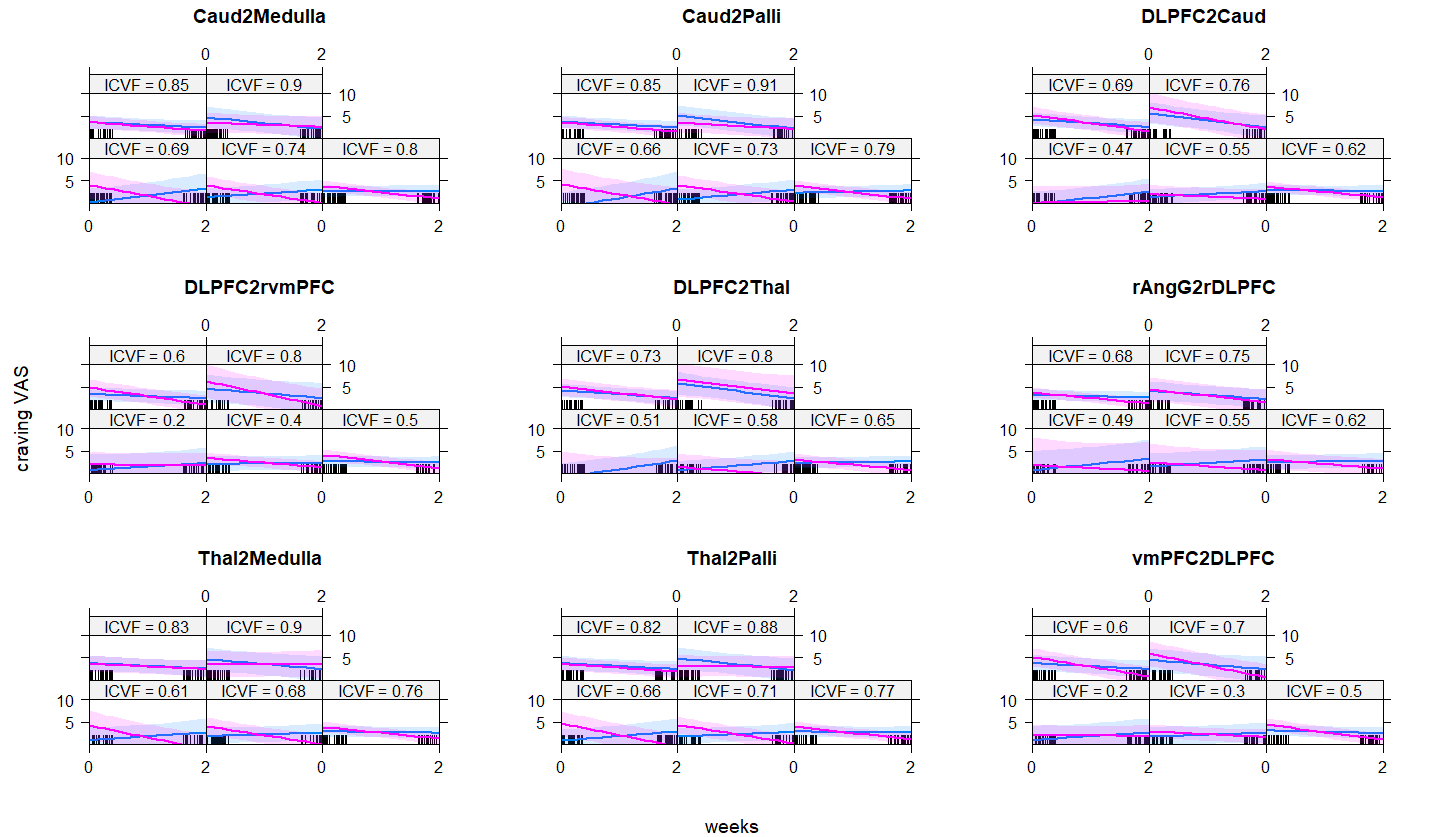

**Supplementary figure 4.** Effect of rTMS and baseline neurite density (ICVF) from frontostriatal circuits’ WM tracts on craving VAS; blue = sham, pink = active.

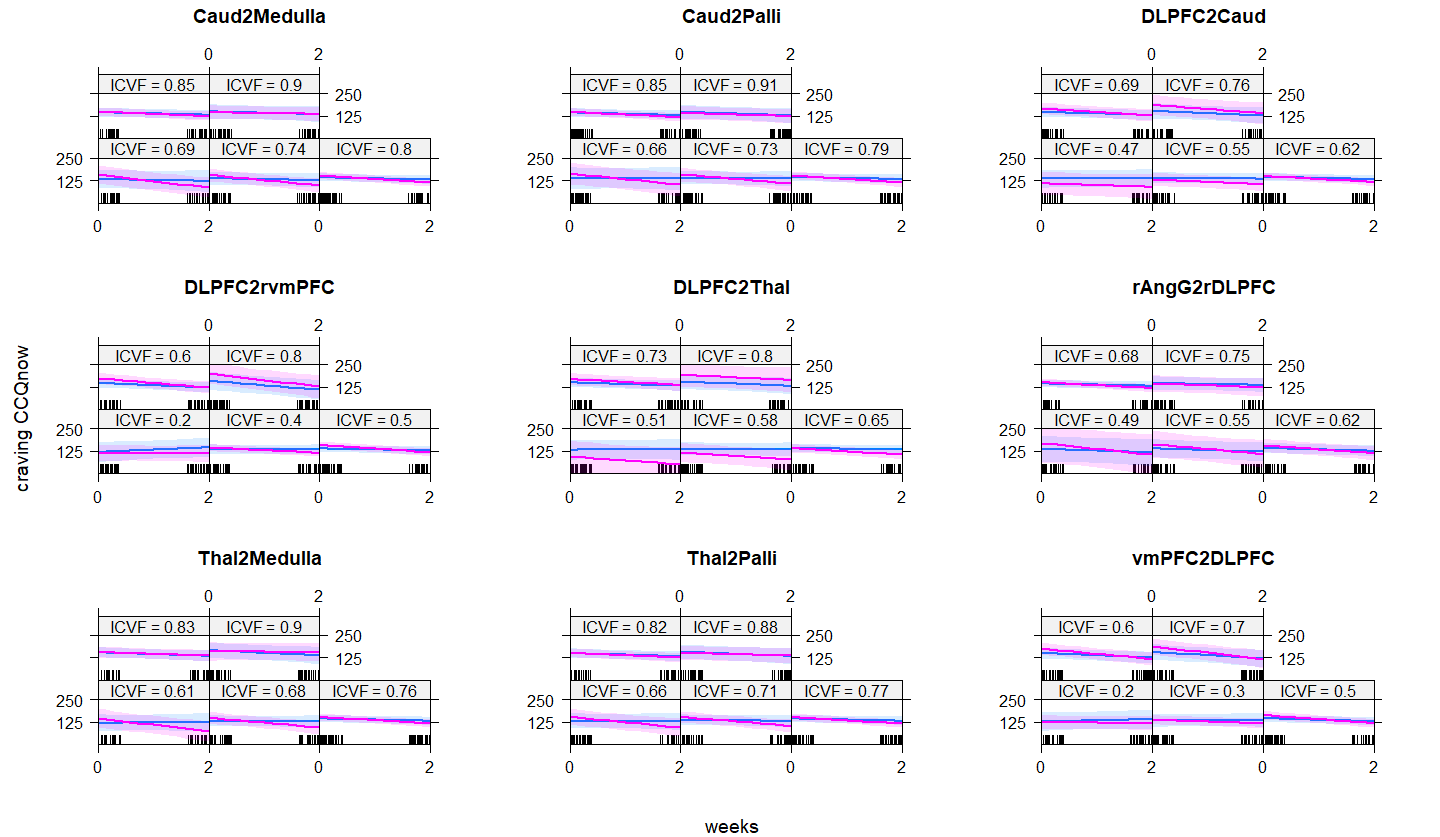

**Supplementary figure 5.** Effect of rTMS and baseline neurite density (ICVF) from frontostriatal circuits’ WM tracts on craving CCQ-now; blue = sham, pink = active.

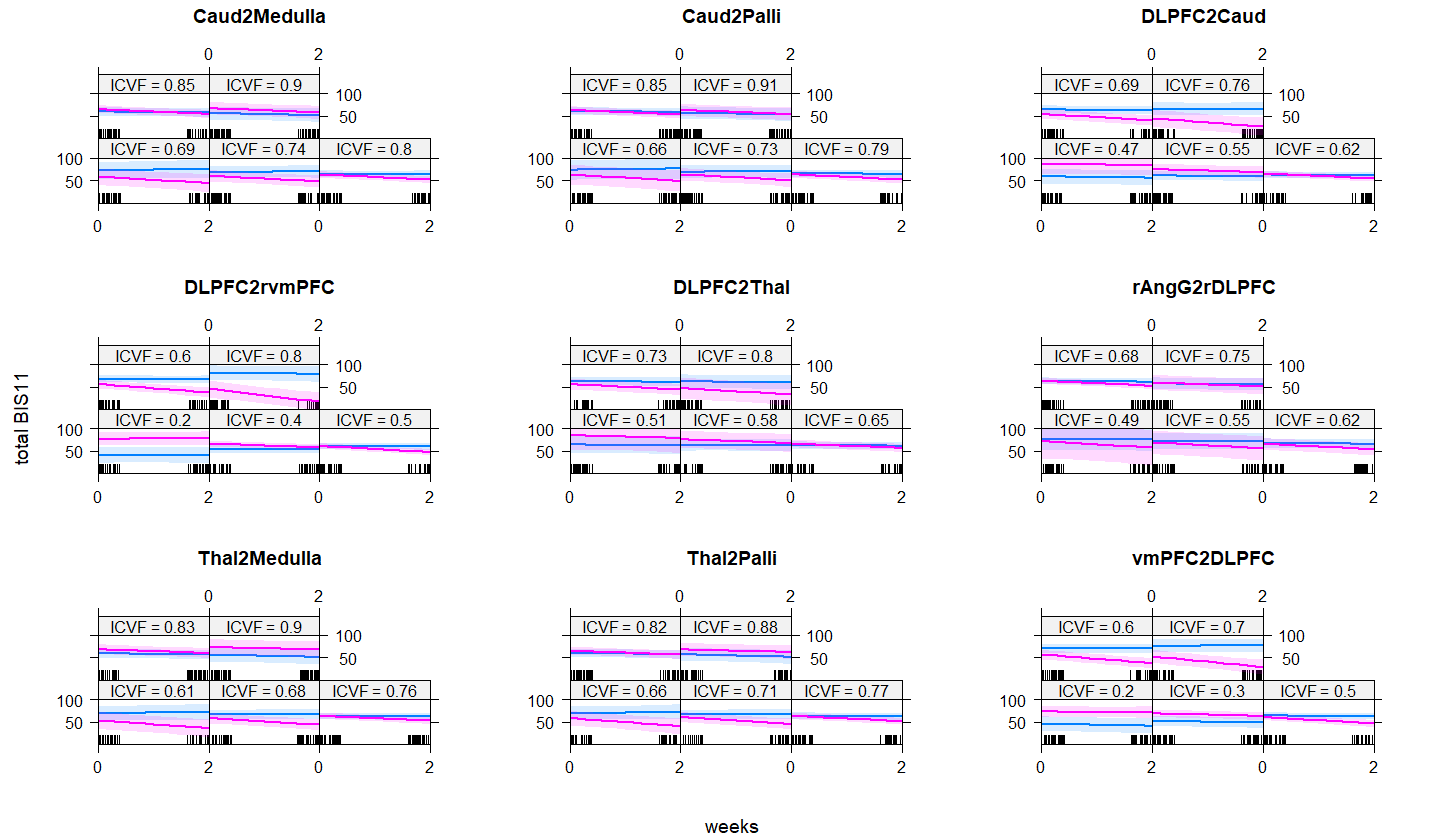

**Supplementary figure 6.** Effect of rTMS and baseline neurite density (ICVF) from frontostriatal circuits’ WM tracts on total BIS11; blue = sham, pink = active.

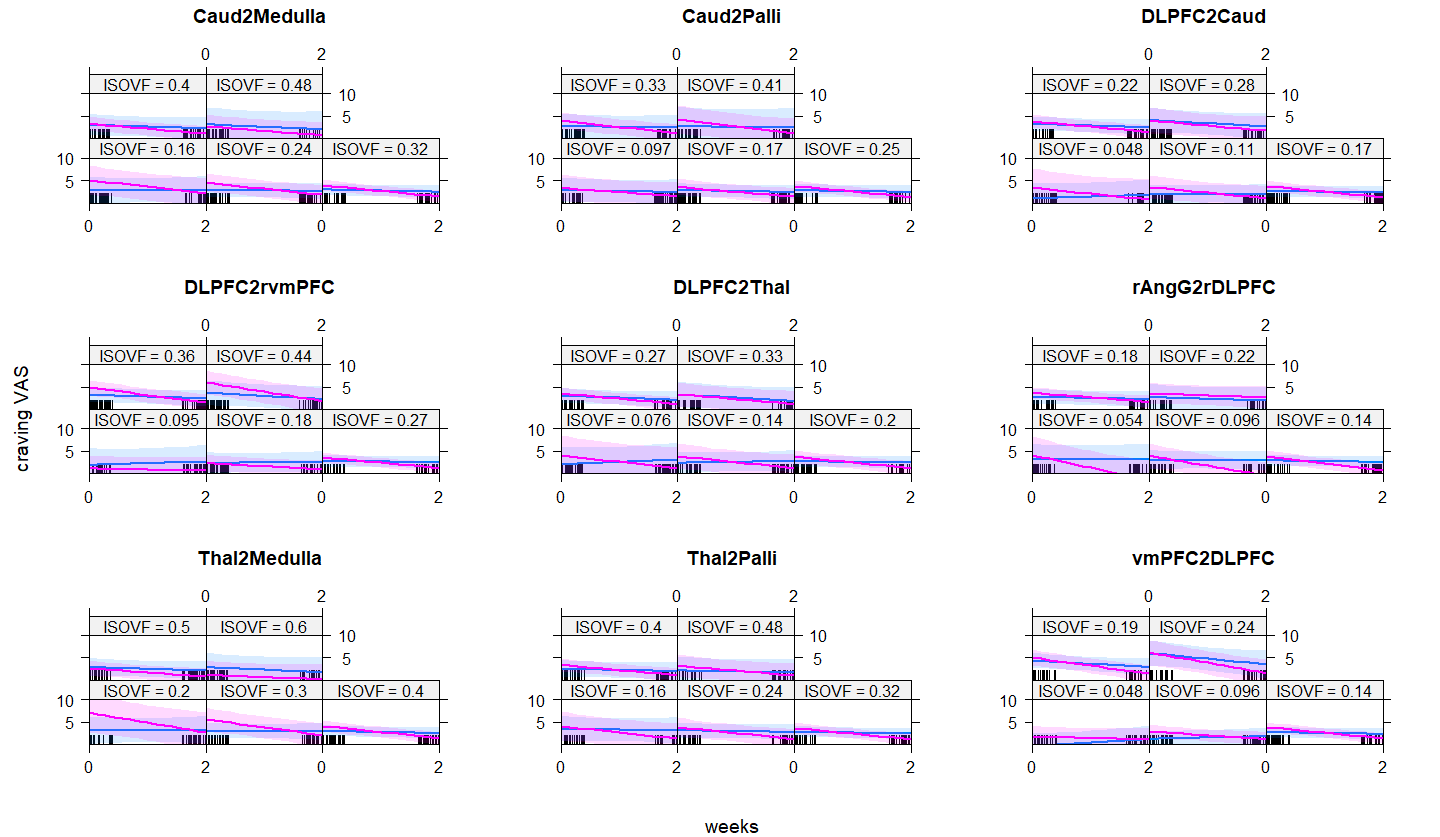

**Supplementary figure 7.** Effect of rTMS and baseline CSF volume fraction (ISOVF) from frontostriatal circuits’ WM tracts on craving VAS; blue = sham, pink = active.

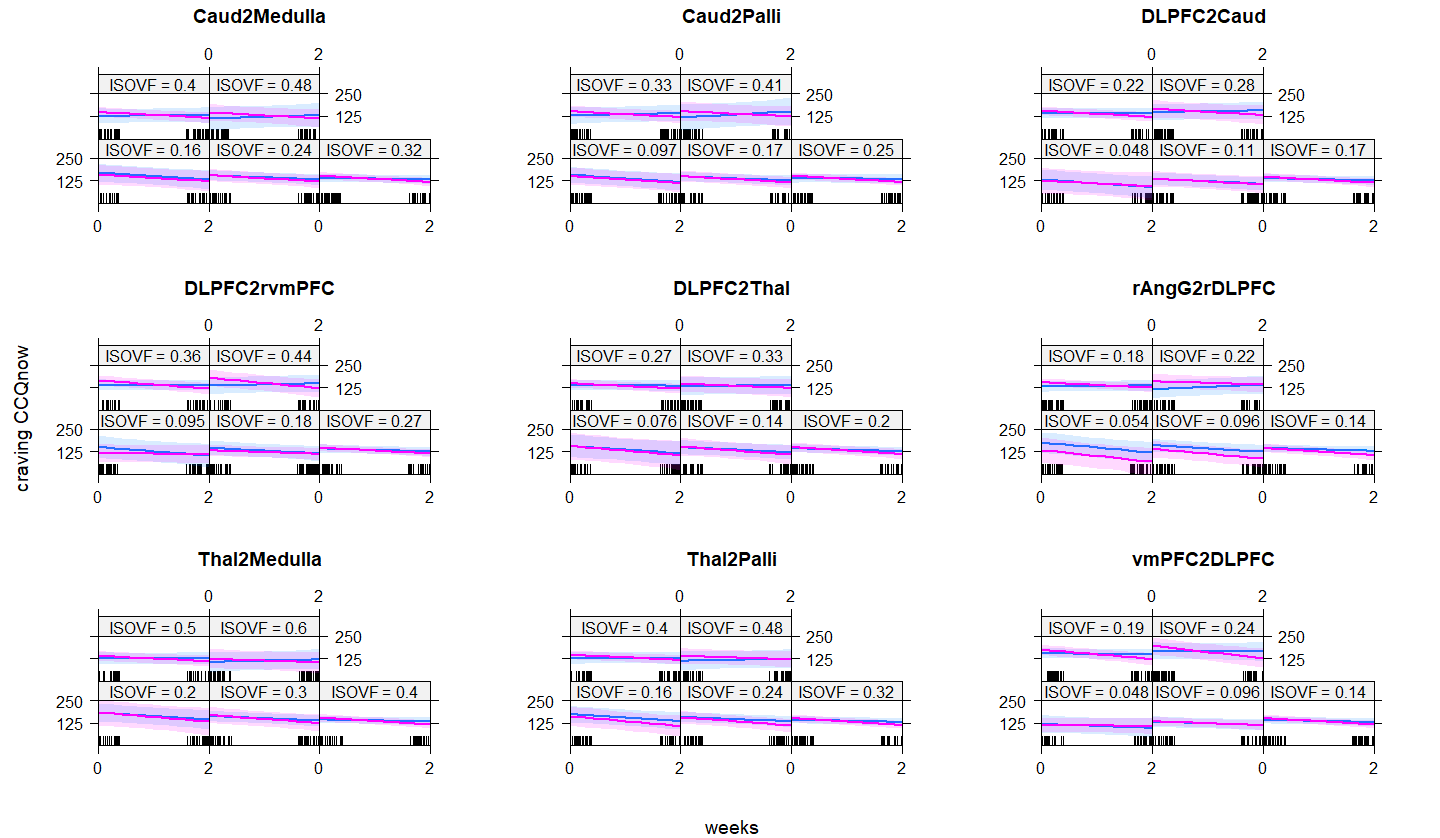

**Supplementary figure 8.** Effect of rTMS and CSF volume fraction (ISOVF) from frontostriatal circuits’ WM tracts on craving CCQ-now; blue = sham, pink = active.

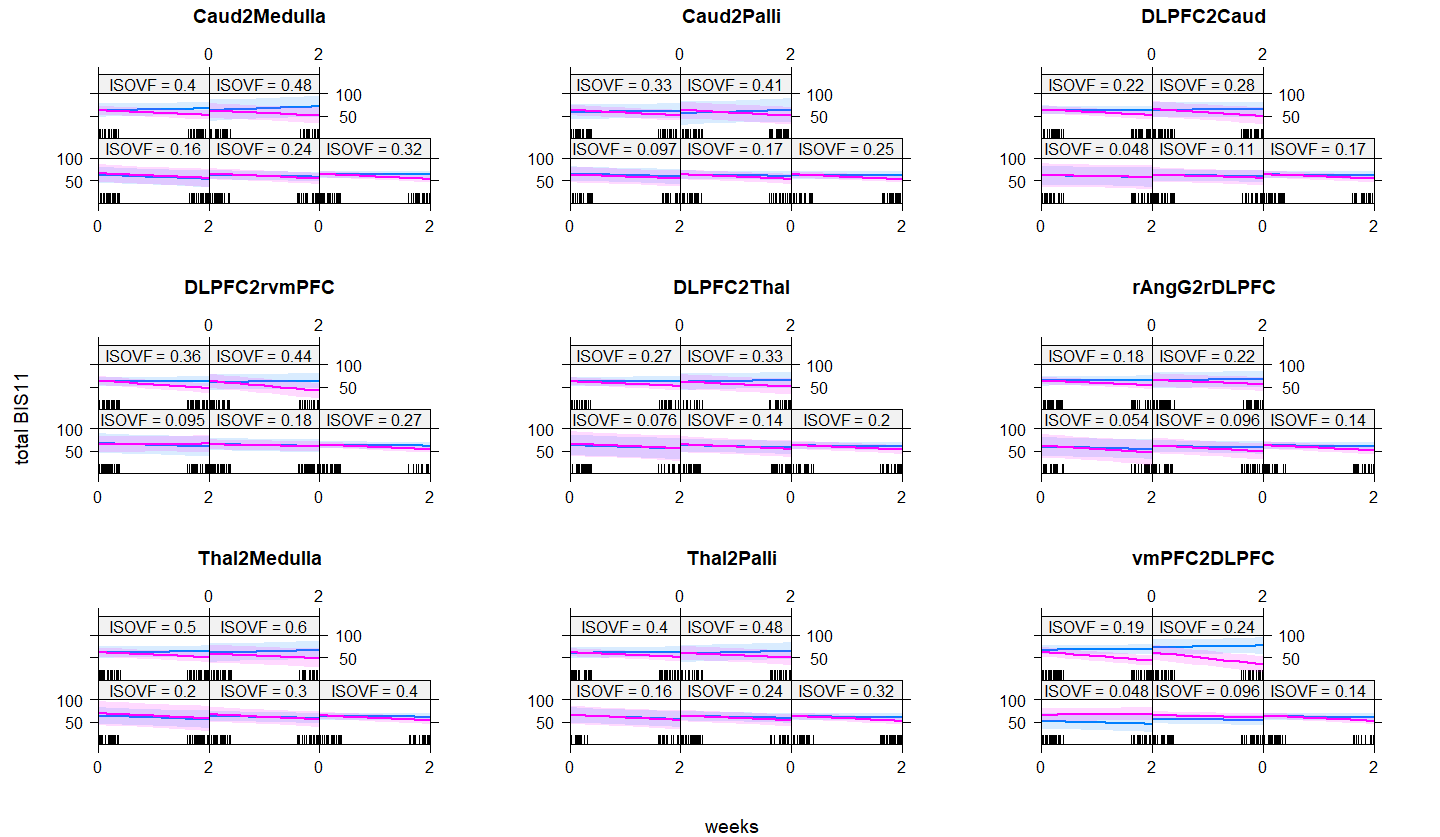

**Supplementary figure 9.** Effect of rTMS and baseline CSF volume fraction (ISOVF) from frontostriatal circuits’ WM tracts on total BIS11.

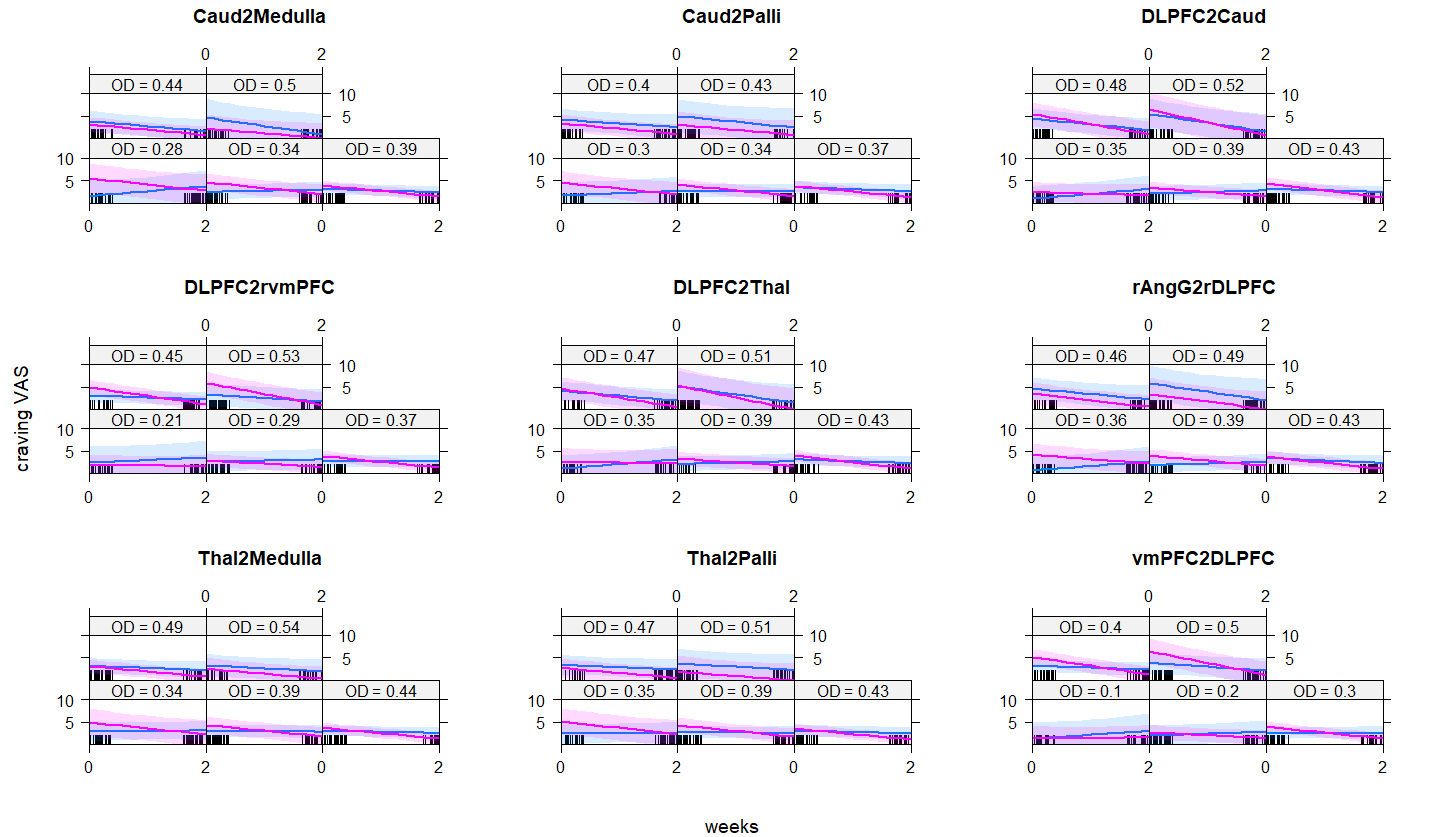

**Supplementary figure 10.** Effect of rTMS and baseline orientation dispersion (OD) from frontostriatal circuits’ WM tracts on craving VAS; blue = sham, pink = active.

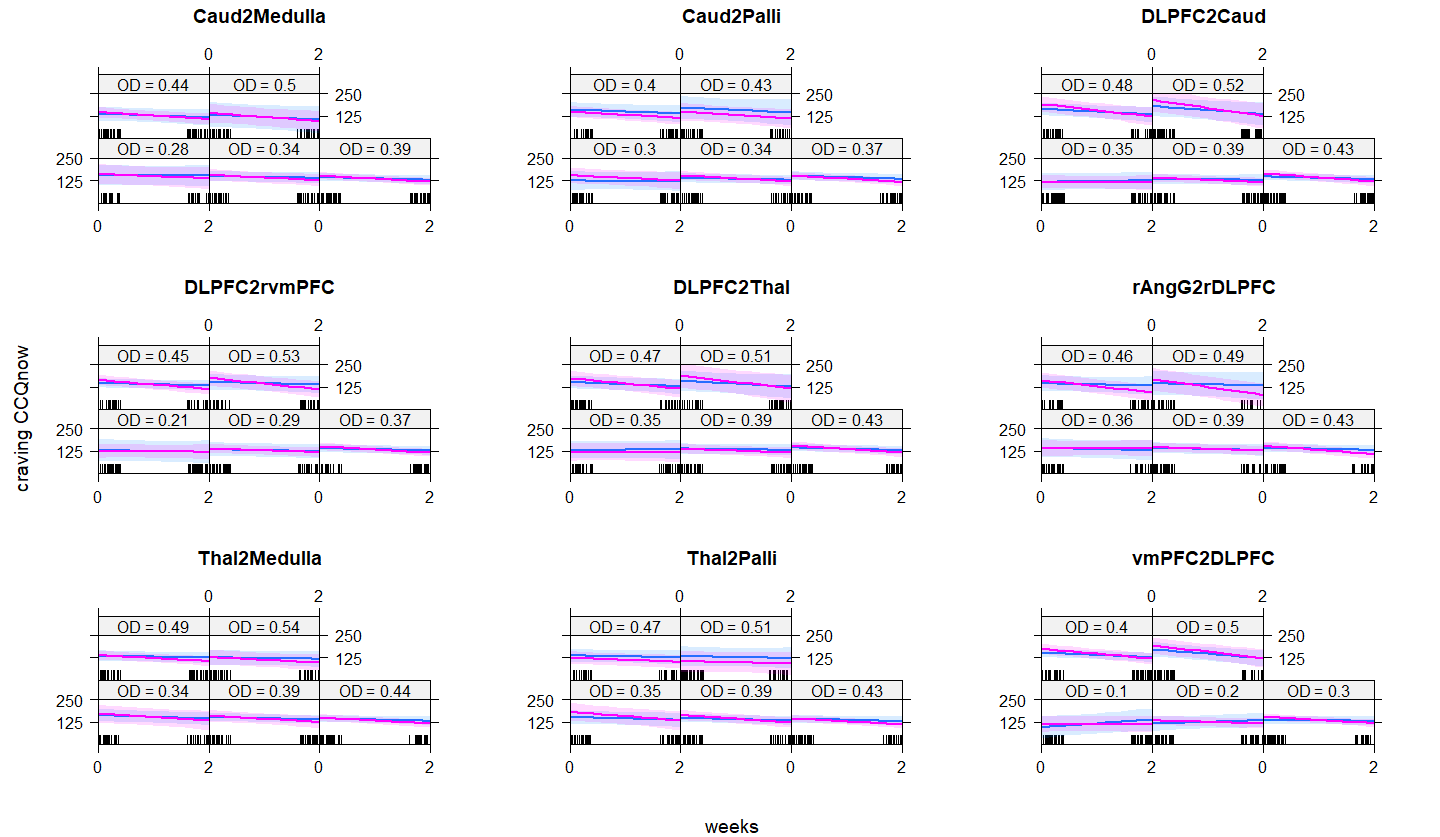

**Supplementary figure 11.** Effect of rTMS and baseline orientation dispersion (OD) from frontostriatal circuits’ WM tracts on craving CCQ-now; blue = sham, pink = active.

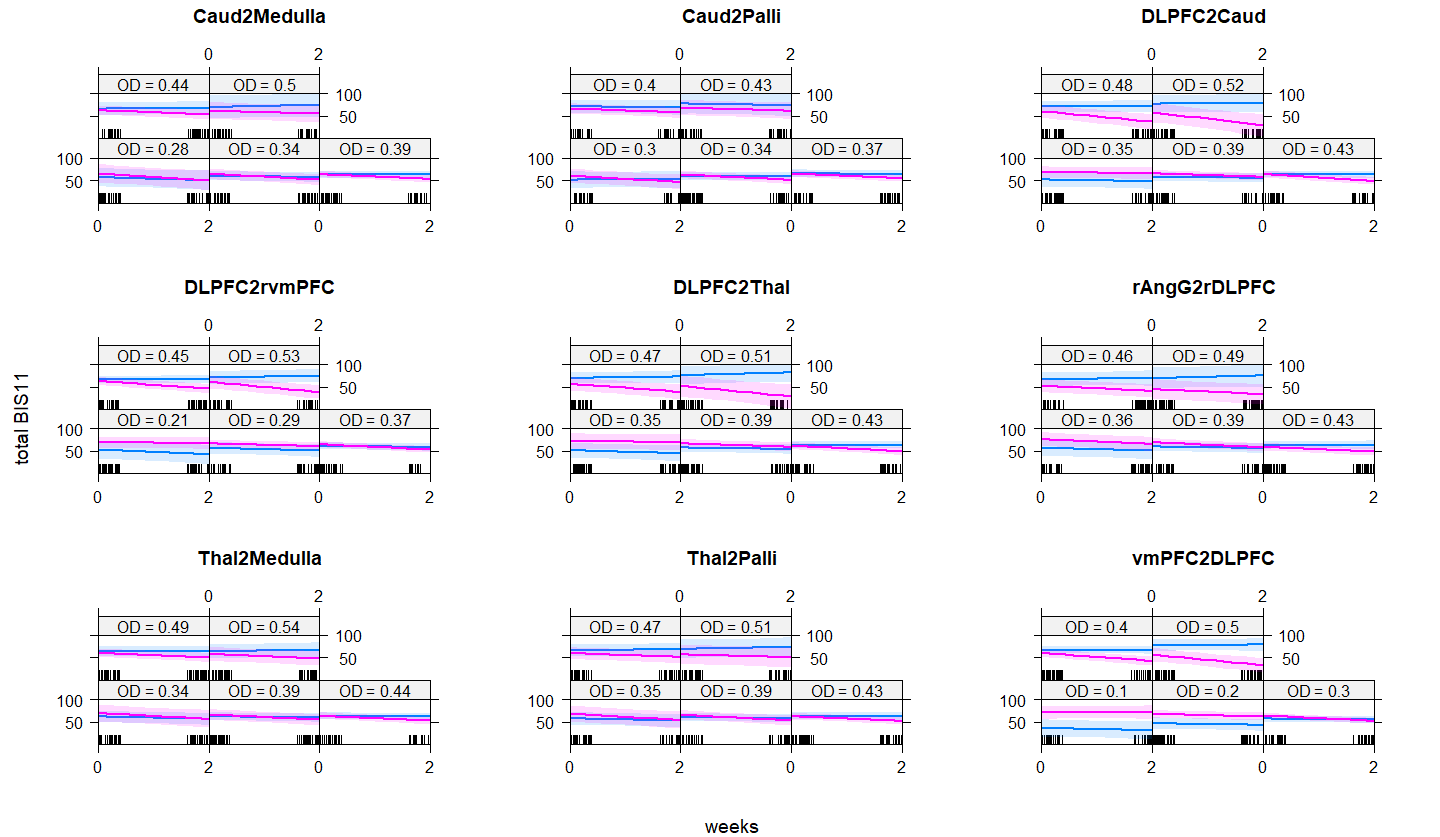

**Supplementary figure 12.** Effect of rTMS and baseline orientation dispersion (OD) from frontostriatal circuits’ white matter tracts on total BIS11; blue = sham, pink = active.
